# Stretch-activated ion channel TMEM63B associates with developmental and epileptic encephalopathies and progressive neurodegeneration

**DOI:** 10.1101/2022.11.22.22282283

**Authors:** Annalisa Vetro, Simona Balestrini, Cristiana Pelorosso, Alessio Masi, Sophie Hambleton, Emanuela Argilli, Valerio Conti, Simone Giubbolini, Rebekah Barrick, Gaber Bergant, Karin Writzl, Emilia K. Bijlsma, Theresa Brunet, Pilar Cacheiro, Davide Mei, Anita Devlin, Mariëtte J.V. Hoffer, Keren Machol, Guido Mannaioni, Masamune Sakamoto, Manoj P. Menezes, Thomas Courtin, Elliott Sherr, Riccardo Parra, Ruth Richardson, Tony Roscioli, Marcello Scala, Celina von Stülpnagel, Damian Smedley, TMEM63B collaborators, The Genomics England Research Consortium, Annalaura Torella, Jun Tohyama, Reiko Koichihara, Keisuke Hamada, Kazuhiro Ogata, Takashi Suzuki, Atsushi Sugie, Jasper J. van der Smagt, Koen van Gassen, Stephanie Valence, Emma Vittery, Mitsuhiro Kato, Naomichi Matsumoto, Gian Michele Ratto, Renzo Guerrini

## Abstract

By converting physical forces into electrical signals or triggering intracellular cascades, stretch-activated ion channels (SACs) allow the cell to respond to osmotic and mechanical stress. Knowledge of the pathophysiological mechanisms underlying associations of SACs with human disease is limited. Here we describe 16 unrelated patients, with severe early onset developmental and epileptic encephalopathy (DEE), intellectual disability, and severe motor and cortical visual impairment, associated with progressive neurodegenerative brain changes, carrying ten distinct *de novo* variants of *TMEM63B*, encoding for a highly conserved SAC. Variants were missense, including the recurrent V44M in 7/16 patients, or in-frame, and affected conserved residues located in transmembrane regions of the protein. In 12 patients, haematological abnormalities co-occurred, such as macrocytosis and haemolysis, requiring blood transfusions in some. We modelled V44M, R443H, and T481N in transfected Neuro2a cells and demonstrated leak inward cation currents across the mutated channel even in isotonic conditions, while the response to hypo-osmotic challenge was impaired, as were the Ca^2+^ transients generated under hypo-osmotic stimulation. Ectopic expression of the V44M and G580C variants in *Drosophila* resulted in early death. *TMEM63B*-associated DEE represents a novel clinicopathological entity in which altered cation conductivity results in a severe neurological phenotype with progressive brain damage and early onset epilepsy, associated with haematological abnormalities in most patients.

## Introduction

*TMEM63B*, with its two paralogues *TMEM63A* and *C*, belongs to a gene family initially identified as the closest homologues of the OSCA proteins, representing the largest family of stretch-activated ion channels conserved across eukaryotes (1, 2). In plants, members of the OSCA family sense osmotic stress-induced mechanical forces across the plasma membrane and activate a signalling pathway responsible for regulating water transpiration and root growth (1, 3). In mammals, members of the TMEM63 family mediate cation currents in response to osmotic and mechanical stimuli affecting membrane tension (1, 4). This process is crucial for cell volume regulation and viability, as changes in osmolarity may cause water influx or efflux through the plasma membrane, resulting in cell swelling or shrinkage (5).

Other members of the *TMEM63* family have been associated with monogenic disorders, namely “transient infantile hypomyelinating leukodystrophy-19 (HLD19)” (MIM #618688), with developmental delay of variable severity, caused by heterozygous *TMEM63A* variants (6–8), and “autosomal recessive spastic paraplegia-87 (SPC-87)” (MIM #619966), caused by biallelic truncating variants of *TMEM63C* (9). The *TMEM63B* gene still lacks a clear association with human diseases, and it is not yet listed either in the OMIM’s Morbid Map of the Human Genome or in the ClinGen Gene-Disease Validity database (https://www.clinicalgenome.org/). Multiple *Tmem63b* mRNA isoforms have been identified in mice, with post-transcriptional modifications contributing to mRNAs diversity (10). A brain-specific *Tmem63b* isoform, exhibiting alternative splicing of exon 4 and glutamine to arginine change (Q/R) at exon 20 has been shown to regulate Ca^2+^ permeability and osmosensitivity of the channel (10). According to GTEx portal (https://www.gtexportal.org/home/), multiple *TMEM63B* mRNA isoforms also occur in humans, with different tissue specificity.

We identified ten distinct heterozygous *de novo* variants of *TMEM63B* in 16 unrelated patients with early onset developmental and epileptic encephalopathy (DEE), all associated with white matter disease, corpus callosum abnormalities, and variable cortical, cerebellar, and haematological abnormalities.

After determining the most represented brain *TMEM63B* isoform in humans, we tested *in vitro* the effect of selected variants on the protein localisation and function by immunocytochemistry, whole-cell patch clamp, and calcium imaging in transfected Neuro2a cells. We also modelled *in vivo* in *Drosophila* the effects of the ectopic expression of two variants. Our findings indicate that heterozygous variants of *TMEM63B* result in a novel and clinically recognizable DEE syndrome whose pathophysiology relies in altered functional properties of the channel.

## Results

### Clinical findings

Clinical, EEG, and MRI findings of the 16 patients are summarized in Table 1 and Supplementary Table 1. They were all unrelated individuals who exhibited a markedly overlapping DEE phenotype with early onset drug-resistant epilepsy (16/16, 100%), severe developmental delay (16/16, 100%), early generalised hypotonia evolving to spastic quadriparesis (13/16, 81%), nystagmus and central visual impairment (11/16, 69%). Epilepsy onset ranged from birth to 3 years but occurred within the first year in 13/16 (81%) and in the first month of life in 5/16 (31%). A common pattern of the epilepsy phenotype was early onset of focal seizures (10/16, 67%), often manifested as apnoeic episodes in newborns, followed over months by epileptic spasms (5/16, 31%) or by different types of focal and generalised onset seizures (6/16, 38%). Infantile epileptic spasms, which were the initial manifestation of epilepsy in three additional patients (3/16, 19%), had therefore been present in 9/16 patients (56%). In two remaining patients (2/16, 13%), onset was between age 2-3 years with focal seizures with impaired awareness. Epilepsy was severe at onset in all patients, with episodes of status epilepticus in three (3/16, 19%). At last follow-up, five patients (5/16, 31%) had no longer severe epilepsy, including three who had experienced prolonged seizure freedom on medication (3/16, 19%). EEGs showed slow background activity with bilateral independent or multifocal epileptiform discharges in most patients.

**Table 1.** Clinical features of the 16 patients with *TMEM63B* variants.

| **Patient number/gender** | **TMEM63B variant (cDNA and protein)/abbreviated** | **Age at last follow-up/death** | **Type of seizure at onset** | **Seizure types/severity during follow-up** | **Treatment ever tried** | **EEG** | **Brain magnetic resonance imaging** | **Clinical neurological phenotype** | **Haematological findings** |
| --- | --- | --- | --- | --- | --- | --- | --- | --- | --- |
| **1/M** | c.130G>A p.(Val44 Met)/V44M | 6-10y | Infantile spasms | Spasms, myoclonic, focal with impaired awareness/daily | CZP, ESM, PB, VGB | Slow background, bilateral independent discharges; epileptic spasms and myoclonic szs | 0-5y: thin CC, colpocephaly, abnormal myelination, dysmorphic lateral ventricles, enlarged extracerebral spaces, progressive mild cerebellar atrophy, and watershed areas WM abnormality | Threatened preterm labour at 35w, global profound DD, generalised hypotonia, plagiocephaly, nystagmus, dysphagia (PEG 0-5y), dyskinesias | Mild anaemia |
| **2/M** | c.1298G>A p.(Arg433 His)/R433H | 6-10y | Focal | Bilateral independent focal motor with impaired awareness, focal to bilateral tonic-clonic/weekly | CBZ, CLB, CZP, LEV, LCM, MDZ, PB, PHT, STP, TPM | Slow background, multifocal discharges; focal szs recorded | 6-10y: thin CC, multifocal WM abnormalities, ventricular asymmetry, progressive cerebellar atrophy | Global profound DD, ataxic gait, lower limb hypertonia, nystagmus | Mild abnormalities of RBCt, MCV, MCH |
| **3/M** | c.1442C>A p.(Thr481 Asn)/T481N | 11-15y | Focal motor with asymmetric posturing | Focal with impaired awareness and asymmetric posturing, focal to bilateral tonic-clonic/weekly | CBZ, CLB, PB, PER, STP, VGB, VNS | Slow background, focal epileptiform discharges | 0-5y: thin CC, posterior predominant multifocal WM abnormalities, dysmorphic asymmetric lateral ventricles, enlarged cortical sulci | Global profound DD, spastic asymmetric quadriparesis, severe cortical visual impairment | None |
| **4/M #** | c.130G>A p.(Val44 Met)/V44M | 11-15y/deceased at 11-15y (pneumonia) | Focal | Focal with impaired awareness; spasms and focal to bilateral tonic-clonic/daily. Single episode of SE | CBZ, CLB, KD, LTG, LEV, PRED, RFM, VGB, VPA, VNS | Slow background, bilateral independent or multifocal discharges; epileptic spasms recorded | 0-5y, 6-10y: thin CC, multifocal WM abnormalities, dysmorphic enlarged lateral ventricles, mild cortical and cerebellar atrophy, progressive trabecular bone thickening | Global profound DD, wheelchair-bound, cortical visual impairment, by 10y knee fixed flexion contractures, dysphagia (PEG 0-5y) | Severe macrocytic anaemia transfusion-dependent |
| **5/M** | c.130G>A<br>p.(Val44<br>Met)/V44<br>M | 6-10y | Apnoeic | Epileptic spasms;<br>focal hyperkinetic<br>motor with<br>impaired<br>awareness/sz-free | CBZ(+), CLB,<br>LEV, PB (+),<br>TPM (+) | Slow background,<br>bilateral independent<br>or multifocal<br>discharges; focal szs<br>recorded | 0-5y: thin CC, abnormal<br>myelination, dysmorphic<br>asymmetric lateral ventricles,<br>enlarged extracerebral spaces,<br>progressive posteriorly<br>predominant WM abnormality<br>and cerebellar atrophy | Global profound DD,<br>severe cortical visual<br>impairment,<br>nystagmus, spastic<br>quadriparesis,<br>dysphagia (PEG 0-5y) | Jaundice at<br>birth |
| **6/F #** | c.130G>A<br>p.(Val44<br>Met)/V44<br>M | 0-5y/<br>deceased<br>at 0-5y<br>(pneumonia in<br>progressive<br>respiratory<br>failure) | Apnoeic | Focal onset with<br>impaired<br>awareness, focal<br>to bilateral tonic-clonic, epileptic<br>spasms/daily | Alimemazine,<br>CBZ, CLB, PB<br>(+/-),<br>paraldehyde,<br>PRED, TPM,<br>VGB, VPA,<br>VNS(+), biotin,<br>pyridoxine | Slow background,<br>multifocal or<br>generalised discharges<br>with burst<br>suppression; focal szs<br>recorded | 0-5y: thin CC, enlarged<br>extracerebral spaces, diffusely<br>abnormal myelination | Global profound DD,<br>quadriparesis with<br>generalised hypotonia,<br>cortical visual<br>impairment,<br>nystagmus, dysphagia<br>(PEG 0-5y) | Macrocytic<br>anaemia,<br>transfusion-dependent |
| **7/M #** | c.130G>A<br>p.(Val44<br>Met)/V44<br>M | 0-5y | Apnoeic | Stiffening<br>episodes;<br>epileptic<br>spasms/sz-free | LEV, steroids,<br>VGB | Normal background,<br>then hypsarrhythmia,<br>slow background,<br>focal discharges | 0-5y: thin CC, widespread WM<br>abnormalities, enlarged<br>dysmorphic lateral ventricles,<br>mild progressive cerebellar<br>atrophy | Global profound DD,<br>asymmetric<br>quadriparesis,<br>generalised hypotonia,<br>cortical visual<br>impairment,<br>nystagmus, dysphagia<br>(no PEG yet) | Scleralicterus |
| **8/F** | c.130G>A<br>p.(Val44<br>Met)/V44<br>M | 16-20y | Infantile<br>spasms | Generalised tonic,<br>focal motor with<br>impaired<br>awareness/daily | ACTH, CBZ,<br>CLB, CZP,<br>ESM, GBP,<br>ivIg, LCM,<br>LEV, PB, PER,<br>PIR, PLP, PRM,<br>STP, TPM,<br>VPA, ZNS | Hypsarrhythmia, then<br>slow background,<br>multifocal<br>epileptiform<br>discharges | 0-5y, 6-10y: thin CC, abnormal<br>myelination, dysmorphic<br>asymmetric lateral ventricles,<br>enlarged extracerebral spaces,<br>progressive widespread WM<br>abnormality, ventricular<br>dilatation, cerebellar atrophy,<br>and trabecular bone thickening | Global profound DD,<br>quadriparesis,<br>dysphagia (PEG 11-15y) | Severe anaemia<br>requiring<br>occasional<br>transfusions |
| **9/M** | c.130G>A<br>p.(Val44<br>Met)/V44<br>M | 11-15y | Febrile<br>seizures | Epileptic spasms,<br>in small clusters/<br>weekly | CZP, VPA | Slow background, no<br>epileptiform<br>discharges in 4 EEG<br>recordings | 0-5y: thin CC with absent<br>splenium and delayed<br>myelination; 6-10y increased<br>signal of WM in watershed areas,<br>generalized decrease of white<br>matter volume | Global profound DD,<br>wheelchair-bound,<br>quadriparesis,<br>pseudobulbar signs,<br>jerky involuntary<br>movements,<br>stereotypical<br>movements and<br>behaviour, dysphagia | Mild anaemia |
|  |  |  |  |  |  |  |  | (PEG 0-5y). Cortical visual impairment, nystagmus |  |
| **10/F** | c.1377C>G<br>p.(Asp459Glu)/D459E | 0-5y | Focal | Bilateral independent focal, epileptic spasms/daily | CBD, KD, LEV, VGB, VNS, ZNS | Slow background, hypsarrhythmia, multifocal discharges; focal szs and epileptic spasms recorded | 0-5y: thin CC, enlarged extracerebral spaces, abnormal myelination more pronounced posteriorly, Rathke cleft cyst | Global profound DD, quadriplegia, generalised hypotonia, cortical blindness with roving eye movements, dysphagia | Mild macrocytic anaemia |
| **11/M** | c.1387G>A<br>p.(Val463Ile)/V463I | 6-10y | Focal | Occasional focal with impaired awareness/during fever | VPA | Normal background, bilateral independent discharges | 6-10y: multifocal, posteriorly predominant WM abnormality | Global severe DD, motor impairment, generalised hypotonia | None |
| **12/F** | c.1424_1426del<br>p.(Ile475del)/I475del | 16-20y | Infantile spasms | Tonic/daily; GTCs /monthly | ACTH, CBD, CLB, CZP, LEV, LTG, OXC, TPM (-), VGB(-), VPA | Hypsarrhythmia, then slow background, bilateral independent or multifocal discharges; epileptic spasms and tonic szs recorded | 0-5y, 6-10y: thin CC, multifocal WM abnormalities, asymmetric dysmorphic lateral ventricles, progressive trabecular bone thickening | Global profound DD, generalised hypotonia, microcephaly, visual impairment, nystagmus, squint, spastic quadriplegia, dysphagia (PEG 11-15y), scoliosis | Severe haemolytic anaemia, transfusion-dependent |
| **13/M** | c.1738G>A<br>p.(Gly580Ser)/G580S | 0-5y | Focal | GTCs/occasional during fever | VPA | Slow background, focal discharges | 0-5y: thin CC, widespread WM abnormality, especially periventricular, asymmetric dysmorphic lateral ventricles, mild cerebellar atrophy | Global profound DD, spastic quadriplegia, axial hypotonia, upper limb dystonia, visual impairment, dysphagia (PEG 0-5y) | None |
| **14/F** | c.1738G>T<br>p.(Gly580Cys)/G580C | 26-30y | Focal | Focal with impaired awareness, GTCs/yearly | CBZ, CZP, PHT(+), VPA, ZNS | Slow background, focal discharges; focal szs recorded | 11-15y, 21-25y, and 26-30y: thin CC, widespread WM abnormality, especially periventricular/posterior, progressive cerebellar atrophy, and trabecular bone thickening | Global moderate DD, cerebellar ataxia, tremor, dysarthria, limited mobility, bipolar disorder | Mild hyperbilirubinaemia |
| **15/F** | c.1979G>C<br>p.(Arg660Thr)/R660T | 21-25y | Focal | Focal with posturing and impaired awareness, recurrent SE/yearly | CBZ(-), LMT(-), OXC, PB(-), TPM, VNS (+) | Background mildly abnormal, diffuse beta activity, multifocal discharges; focal szs recorded. | 0-5y, 6-10y: thin CC, multifocal WM abnormality | Global moderate DD, spastic quadriplegia, ASD | Macrocytic anaemia |
| **16/M** | c.2089T>C<br>p.(Phe697Leu)/F697L | 11-15y | Focal | Focal with eye deviation-jerking, GTCs, recurrent SE/sz-free at last FU | CLB, LTG(-), LEV(-), OXC, PHT, VPA | Normal background; focal discharges | 0-5y, 11-15y: thin CC, abnormal myelination, colpocephaly, dysmorphic lateral ventricles, increase WM signal with posterior/periventricular predominance | Global, severe DD, toe walking, ASD | None |

Abbreviations and symbols: ASD, autism spectrum disorder; CBD, cannabidiol; CBZ, carbamazepine; CC, corpus callosum; CLB, clobazam; CZP, clonazepam; DD, developmental delay; ESM, ethosuximide; F, female; GBP, gabapentin; GTCS, generalized tonic-clonic seizures; ivIg, intravenous Immunoglobulin; KD, ketogenic diet; LCM, lacosamide; LEV, levetiracetam; LTG, lamotrigine ; M, male; MCH, mean corpuscular haemoglobin; MCV, mean corpuscular volume; MDZ, midazolam; NA, not available; OXC, oxcarbazepine; PB, phenobarbital; PEG, percutaneous endoscopic gastrostomy; PER, perampanel; PHT, phenytoin; PIR, piracetam; PLP, pyridoxal 5'-phosphate; PRED, prednisolone; PRM, primidone; RBC, red blood cells; RFM, rufinamide; SE, status epilepticus; STP, stiripentol; sz, seizure; TGP, targeted gene panel; TPM, topiramate; VGB, vigabatrin; VNS, vagus nerve stimulation; VPA, valproic acid; WM, white matter; y, years; ZNS, zonisamide; #, Pts 4, 6 and 7 are mentioned in Cacheiro et al, 2020 (23)(Supplementary Table S8: DDD patient1, 100KGP patient 2, and 100KGP patient 1); treatment ever tried: +, efficacy; +/-, transient efficacy; -, worsening or not tolerated.

All patients (16/16, 100%) had global developmental delay, with moderate to profound intellectual disability and severe motor impairment; only two had developed communicative language skills (2/16, 13%). Severe dysphagia was present in ten patients (10/16, 63%), requiring a percutaneous endoscopic gastrostomy (PEG) insertion in eight (8/16, 50%). In 12 patients (12/16, 75%) clinical history revealed haematological abnormalities resulting in abnormal haemoglobin levels, often with macrocytosis, and signs of haemolysis (hyperbilirubinemia, jaundice, hepatosplenomegaly), with a fluctuating course. In four of the 12 patients (4/12, 33%) with haematological abnormalities, three of whom carrying the recurrent V44M variant, anaemia was severe and required blood transfusions (Pts 4, 6, 8, 12). In Pt 4, peripheral blood film showed abnormally shaped red cells (RBCs) with elliptocytes and increased stomatocytes. Bone marrow biopsy was performed in two patients (Pts 4 and 8, 2/16, 13%) with signs of haemolysis in both, and evidence for haematopoiesis and myelodysplastic syndrome with aplastic anaemia in Pt 8 (Supplementary Figure 1). Growth parameters at birth were abnormal in terms of weight (<3^rd^ percentile in 1/16, 6% and >99^th^ in 1/16, 6%) and length (<3^rd^ pc in 3/16, 19%). Patients with normal birth growth parameters showed subsequent signs of growth failure (<3^rd^ pc, 6/16, 38%). Two patients (2/16, 13%) had died prematurely due to pneumonia (Pts 4 and 6; Table 1). Microcephaly was present in three patients (3/16, 19%), in one of them since birth along with severe haemolytic anaemia (Pt 12).

Prompted by the evidence for progressive hearing loss in *Tmem63b* deleted mice (4), we investigated whether any clinical sign of hearing impairment had occurred, reviewed the results of formal hearing testing in 10/16 (63%) patients, and found no relevant abnormalities.

### Magnetic resonance imaging findings

All patients had at least one brain MRI scan at 1.5 or 3T between ages one week and 28 years, and 11 patients were studied with two or more scans taken six months to 13 years apart (Table 1 and Figure 1). MRI revealed an association of multiregional or widespread white matter signal abnormalities, dysmorphic lateral ventricles, thinning of corpus callosum (16/16, 100%), cerebellar atrophy (8/16, 50%), and atrophy of the cerebral cortex (7/16, 44%).

**Figure 1.**
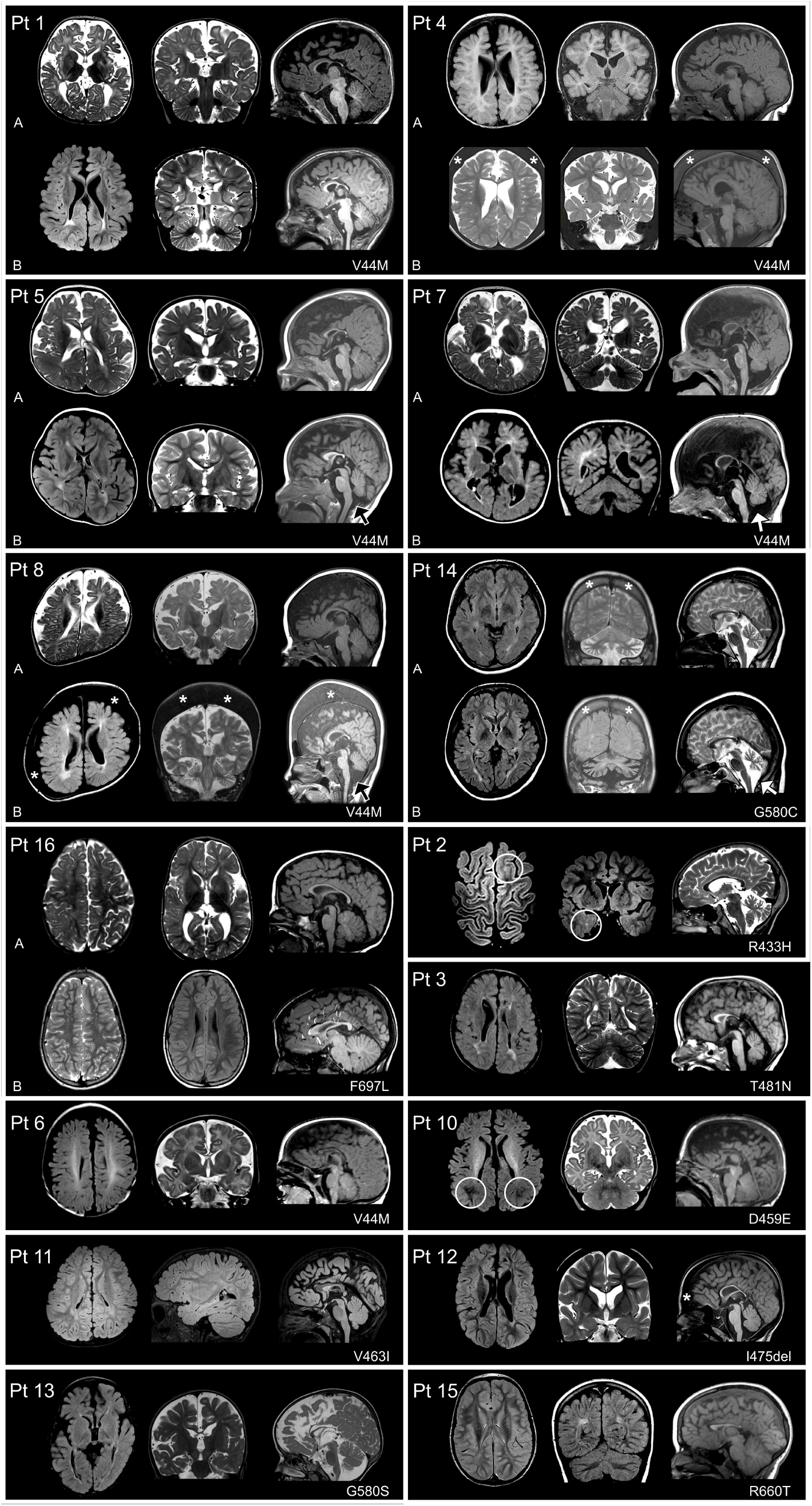
Brain MRI in 15 patients with *TMEM63B* pathogenic variants. Patients’ numbers are shown in the upper left corner. For each patient, the corresponding *TMEM63B* variant is reported in the lower right corner. MRIs of the 10 patients who were imaged at least twice are shown. For Pts 1, 4, 5, 7, 8, 14, and 16 two sets of comparative images are presented, from the initial (A) and follow-up (B) investigation. We show axial (left column), coronal (middle column), and sagittal (right column) images for all patients, except for Pt 16 where two axial (left and middle column) and one sagittal (right column) images are shown. For Pts 2, 3, 6, 10, 11-13, and 15, one set of axial (left column), coronal (middle column), and sagittal (right column) images is presented. Although Pts 2, 12, 15 had serial MRIs, only one set of images is presented as either there were not significant changes or not all sets of images were available. Images were taken at 1.5 to 3T and include T_1_-or T_2_-weighted and FLAIR sequences. Structural abnormalities include a combination of white matter abnormalities, dysmorphic lateral ventricles, thinning of corpus callosum, cortical and cerebellar atrophy that are variably distributed. In Pt 1, at age 0-5 years, the main features include increased extracerebral spaces with enlarged cortical sulci, thin corpus callosum, with consequent colpocephaly, and high signal intensity of the hemispheric white matter, consistent with abnormal myelination. At a later follow-up, enlarged extracerebral spaces and thinning of the corpus callosum are less prominent but there is a definite high signal abnormality of the white matter, with dysmorphic lateral ventricles, and mild atrophy of the cerebellar cortex. In Pt 4, after a first MRI at age 0-5 years, only showing reduced volume of the frontal lobes, a follow-up scan revealed a progressive change in shape of the lateral ventricles and corpus callosum, both revealing white matter suffering, with mild atrophy of the cerebellar cortex. In Pts 5 and 8, changes that occurred from age 0-5 years over an interval of 1 to 9 years are comparable to those observed in Pt 1. The abnormal ventricular shape causes ventricular asymmetry in these patients. Cerebellar atrophy is indicated by black arrows. In Pt 7 a second MRI at age 0-5 years confirms the severe white matter abnormality, with dilated ventricles and thin corpus callosum already visible in the first images, and revels mild progressive cerebellar atrophy (white arrow). In Pt 14, who was imaged twice 6-10 years apart, MRI images show as white matter changes and cerebellar atrophy continued to progress in adulthood, after age 26-30 (white arrow). In Pt 16, only minor changes occurred between age 0-5 and 11-15 years, with no clear worsening. At 0-5 years, there were areas of abnormal myelination, especially on the right hemisphere and a thin corpus callosum with colpocephaly. At age 11-15, the corpus callosum remained thin but because of maturation processes was thicker than before and ventricular dilatation less prominent, with mild peritrigonal high signal intensity abnormality. In Pt 2, brain MRI, taken at 6-10 years, shows bilateral patchy areas of white matter abnormalities highlighted by a circle in the left frontal and right temporal regions. This finding, in association with focal seizures had initially raised the suspicion of areas of focal cortical dysplasia. Additional findings include ventricular asymmetry, thin corpus callosum and cerebellar atrophy. Cerebellar atrophy had progressed when compared with previous imaging taken at age 6-10 years (images not shown). In Pt 3, who was imaged at 0-5 years, there are multifocal high signal white matter changes with dysmorphic and asymmetric lateral ventricles, thin corpus callosum, and enlarged cortical sulci. Pts 6 and 10, both imaged at age 0-5 years, exhibited increased extracerebral spaces with enlarged sulci, thin corpus callosum, and reduced signal intensity of the white matter, consistent with abnormal myelination. The abnormal myelination is more obvious posteriorly in Pt 10 (white circles). In Pt 11, at age 6-10 years, there were only multifocal areas of white matter abnormalities without atrophic changes. Pts 12, 13, and 15 imaged between 0-5 and 6-10 years, show similar findings, slightly varying in severity and including a thin corpus callosum, areas of abnormal signal intensity of the white matter with posterior predominance (present in all), dilated asymmetric ventricles (Pts 12 and 13), and mild cerebellar atrophy (Pts 13). In Pts 4, 8, 12, and 14 thickening of the trabecular (spongy) bone of the skull (indicated by the asterisks) is suggestive of a blood disorder.

All the 11 individuals for whom repeated MRIs were available for comparison (Pts 1, 2, 4, 5, 7, 8, 9, 12, and 14-16), exhibited clear morphological or signal signs resulting from the combination of maturational and disease-related progressive changes. For example, when cerebellar atrophy was present, there had always been evidence of atrophy progression from initial to follow-up imaging (Pts 1, 2, 4, 5, 7, 8, and 14). White matter signal abnormalities (always present) appeared in early scans as severely delayed myelination that was better appreciated using T2-weighted imaging and later evident as diffuse or more circumscribed areas of high signal intensity changes in FLAIR (see Pts 1, 5, 7, and 8 in Figure 1). In the only individual who had received control MRI scans at different adult ages (Pt 14), cerebellar and white matter changes continued to worsen between age 6-10y. Overall, white matter abnormalities, ventricular shape, and thinning of the corpus callosum exhibited similar longitudinal changes. Signs of progression of cortical atrophy were minor, if any, and proportional to overall volume shrinking. MRI also showed progressive thickening of the trabecular (spongy) bone of the skull in five patients (5/16, 25%; Pts 4, 8, 12, and 14 in Figure 1; Pt 9, not shown) four of whom had anaemia (Pts 4, 8, 9, and 12), with a myelodysplastic disorder in three (Pts 4, 8, and 12) while one had experienced episodes of hyperbilirubinemia (Pt 14).

### Genetic findings

In these 16 patients, we identified ten distinct *TMEM63B* variants (Table 1, Figure 2) including nine missense substitutions and one in-frame deletion. None of them was present in publicly available allele frequency databases such as GnomAD and TOPMed, or in our internal dataset, and all were predicted to be damaging by multiple prediction tools (Supplementary Table 2). Available gene constraint scores from GnomAD indicate that *TMEM63B* is globally highly intolerant to both loss-of-function (LoF; pLI score = 1.00; observed/expected, o/e LoF = 0.07) and missense (Z score = 4.22; o/e missense = 0.475) variants. In addition, all variants identified in our patients were in regions of the protein predicted to be intolerant to missense variations according to region-level metrics (Figure 2B, Supplementary Figure 2) and all involved amino acids highly conserved among the vertebrate orthologues of the protein (Figure 2C, Supplementary Table 2), as well as the paralogues *TMEM63A* and *TMEM63C* (Supplementary Figure 3).

**Figure 2.**
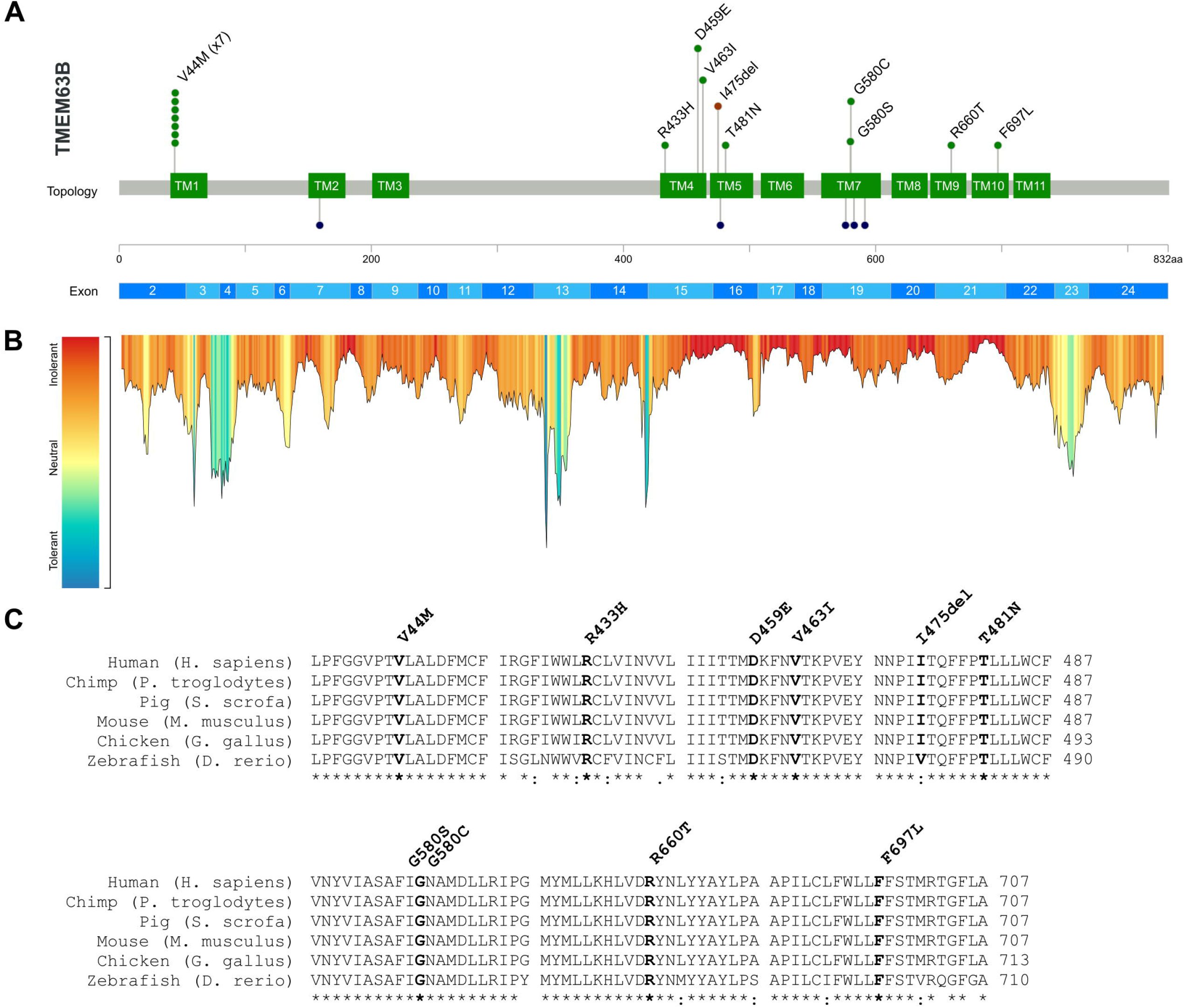
Genetic results. (A) The lollipop diagram shows the distribution of the TMEM63B variants observed in our cohort on the linear protein map and relative to the *TMEM63B* exons (top, based on NM_018426.3 sequence). The variants are represented as green (missense substitutions) or brown (in-frame deletion) dots and all map in the transmembrane helices TM1, TM4-7, and TM9-10 (green boxes). The V44M is recurrent in seven patients, as illustrated by the number of green dots. At the bottom of the diagram, the dark blue dots represent the residues affected by missense variants in *TMEM63A* (five variants in six unrelated patients: G168E, I462N, G553V, Y559H, G567S) (6–8). (B) Tolerance landscape plot of the TMEM63B protein provided by the MetaDome web server (https://stuart.radboudumc.nl/metadome/). The tool identifies regions of low tolerance to missense variations based on the local non-synonymous over synonymous variants ratio from GnomAD (46). All variants in our cohort are contained in intolerant/highly intolerant regions (in red) of the landscape. (C) Multiple sequence alignment shows the protein sequence of the human TMEM63B protein (NP_060896.1) and of its orthologues in five different vertebrate species (*Pan troglodytes, Sus scrofa, Mus musculus, Gallus gallus, Danio rerio*), with the mutated residues in bold. The details of the TMEM63B variants in the cohort are displayed above the alignments. The asterisk below the sequence indicates positions which have a single, fully conserved residue between all the input sequences, the colon indicates conservation between groups of strongly similar properties, and the period indicates conservation between groups of weakly similar properties.

We noticed a complementary linear distribution on the protein sequence of the *TMEM63B* variants in our cohort versus the gene variants in the reference population from GnomAD. The latter were enriched in exons 3-14 and 22-24 and substantially depleted in exons 15-21, where our variants clustered (Figure 2B, Supplementary Figure 2). This observation, supported by the local constraint metrics provided by multiple tools, suggests an increased selective pressure in the region of the gene corresponding to the seven transmembrane domain region RSN1_7TM (PF02714). This domain is conserved among osmosensitive calcium-permeable cation channels (11).

The V44M variant was recurrent in seven unrelated patients (Pts 1, 4-9) and two different changes affected the same residues in two patients (G580S in Pt 13 and G580C in Pt 14). Eight of the affected residues are fully conserved from human to zebrafish (V44, R443, D459, V463, T481, G580, R660, and F697), and one is fully conserved up to chicken (I475). In addition, six out of nine affected residues are fully conserved among all the three paralogues’ sequences (V44, D459, V463, G580, R660, and F697), and three (R443, T481 and I475) among two of the three paralogues, maintaining strongly similar properties in the third (Supplementary Figure 3).

The *TMEM63B* variant occurred *de novo* in the 15 patients for whom parental DNA was tested. Patient 10 was born through gamete-donation and we could not analyse parental DNA.

Based on both *in silico* analyses using sequence data and the functional studies described below, we interpreted all variants as detrimental for the protein function.

### Structural considerations

Since crystallographic data are not yet available for TMEM63B, we can only access information about the protein topology based on its partial homology with the plants’ OSCAs, for a subset of whom crystallographic data are available (2, 12, 13). As the percentage of identity in protein sequence between mammals TMEM63A-C proteins and OSCAs is around 20% (1), mapping our variants on these structures would be inaccurate. Aware of the limitations of prediction tools in regions of low homology, we mapped our variants on the structure exhibiting more homogenous resolution across the protein, including both the transmembrane helices and the intracellular domains (Figure 3, Supplementary Figure 4). In our model, 81% of residues had >90% confidence, including all those affected by variants in our cohort, all affecting a predicted transmembrane (TM) helix (Figure 2A and 3A, B). Five variants fell in TM4 (R433H, D459E, and V463I) or TM5 (I475del and T481N) (Figure 3A, B, blue helices), whose tilting and rearrangement upon osmotic stimulus plays a role in channel opening in *Oryza sativa* OSCA1.2 protein (13). The remaining variants mapped in TM1 (V44M), TM7 (G580S and G580C), TM8 (R660T), and TM9 (F697L). All but the recurrent V44M variant are in the Pfam-classified domain RSN1_7TM (11) and six of them affect the predicted pore-forming TM4-TM8 helices (2).

**Figure 3.**
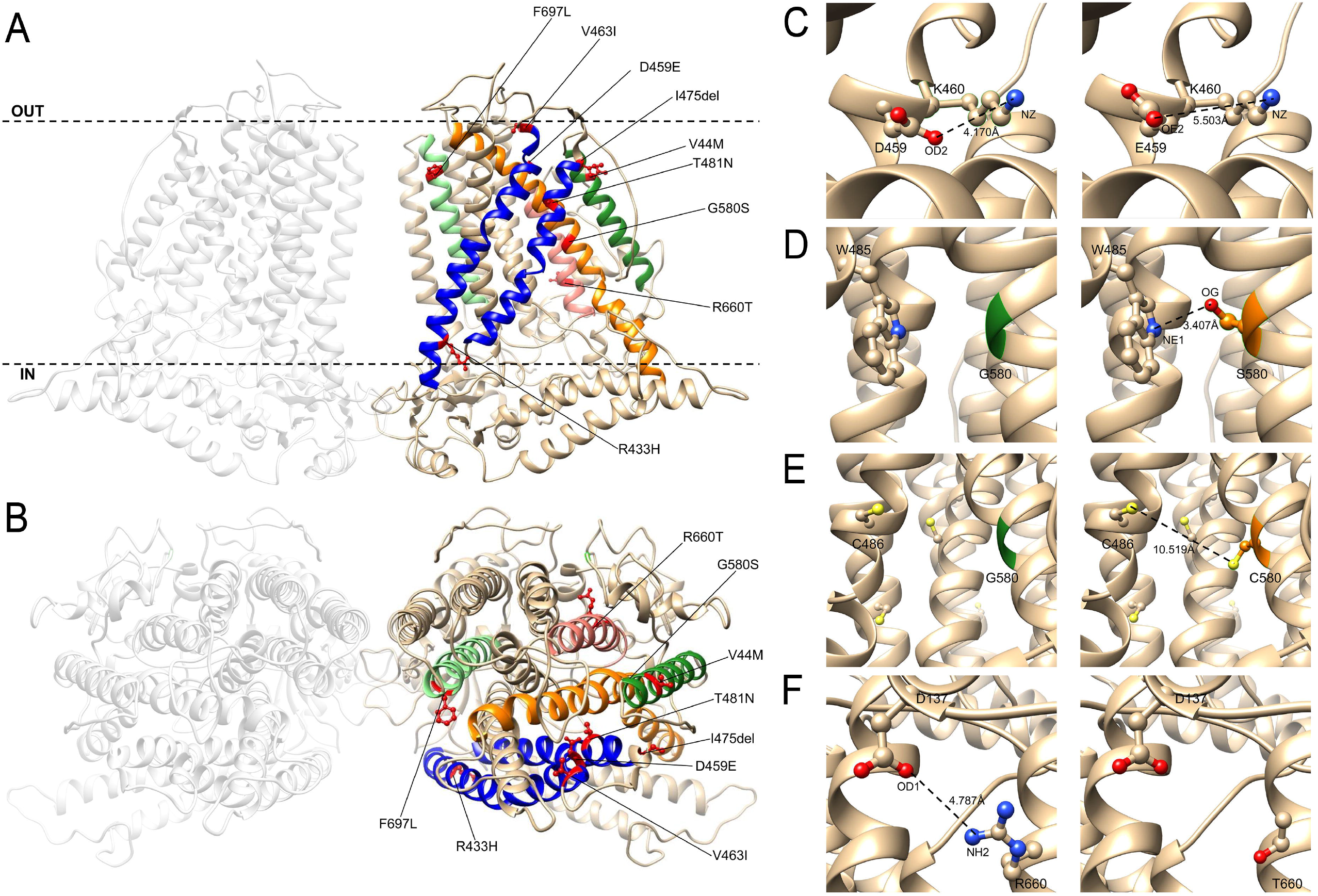
Structural consideration of TMEM63B pathogenic variants. View of the predicted tridimensional protein structure of TMEM63B from the membrane plane (A) and the extracellular side (B). All the variants in our cohort map into a transmembrane (TM) helix: V44M in TM1 (dark green helix), R433H, D459E, V463I, I475del, and T481N in TM4 and TM5 (blue helices), G580S in TM7 (orange helix), R660T in TM8 (pink helix), and F697L in TM9 (light green helix). Dotted line in (A) indicate the plasma membrane, OUT the extracellular side and IN the intracellular (cytoplasmic) side. Details of selected variants are provided in the inlets. (C) Predicted structural change induced by the D459E substitution. The OD2 atom of Asp459 is predicted to form a buried salt bridge with NZ atom of Lys460 (K460). The substitution of an aspartic acid with a glutamic acid at position 459 increases the distance between the NZ atom of Lys460 and the closer oxygen atom (OE2) available to make a salt bridge, breaking this bound. (D) Predicted structural change induced by the G580S substitution. The substitution of a glycine (green) with a bulkier amino acid (serine, orange) changes the RSA of the amino acid at position 580 (5.9% to 3.8%). In addition, OG atom of Ser580 might form a salt bridge with NE1 atom of Trp485 (W485) and help in stabilizing the structure of the pore. (E) Predicted structural change induced by the G580C substitution. The substitution of a Glycine (green) with a bulkier amino acid (Cysteine, orange) changes the RSA of the amino acid at position 580 (5.9% to 3.7%). Although the substitution introduces an amino acid with a free SH group which can make disulphide bonds with other amino acids with free SH groups (depicted as yellow spheres), the distance between C580 and the closer amino acid with a free SH group (C486, 10.519Å) is too big to allow the making of such type of bond. (F) Predicted structural change induced by the R660T substitution. The substitution of a buried charged residue (Arginine) with an uncharged residue (Threonine) at position 660 disrupts a salt bridge formed by NH2 atom of Arg660 and Asp137 (D137).

The V44M (Pts 1, and 4-9) lies in a region with limited sequence homology between OSCAs and TMEM63A-C, and for which available OSCAs structures have low resolution (2, 12). The Valine to Methionine substitution involves two amino acids with similar properties (Grantham distance 21; scale 0-215) and mass. The TM1 helix is located on the external surface of the channel and is not directly involved in the predicted pore, but seems to be rather involved, together with TM7, in the sensitivity to membrane stretching (2, 12) (Figure 2).

In a FoldX-based model, Met44 is predicted to make van der Waals contacts with Gln477, Phe478, and Thr481, all mapping on the TM4 helix (Supplementary Figure 5). Since the free energy change associated with V44M is negative (−1.46, calculated by FoldX), we hypothesize that this substitution may stabilize the protein structure.

We estimated evolutionary conservation and role of the mutated residues by ConSurf, which confirmed that all changes affected highly conserved residues with predicted functional and/or structural role (Supplementary Figure 6; Supplementary Table 3).

We also evaluated the structural impact of the nine distinct missense variants in our cohort using the Missense3D tool (14) and found that 8/9 may affect the protein structure, although limitedly, via changes in cavities volume (7/8), residues accessibility (3/8), and breakage of non-covalent bonds (2/8) (Supplementary Table 3). The D459E substitution (Pt 10) disrupts a salt bridge formed between OD2 atom of Asp459 and NZ atom of Lys460 (distances: 4.168 Å between OD2 atom of Asp450 and NZ atom of Lys460, 5.503Å between OE2 atom of Glu450 and NZ atom of Lys460) (Figure 3C). The G580S substitution (Pt 13) replaces a buried Glycine, with a residue solvent accessibility (RSA) of 5.9%, with a buried Serine with an RSA of 3.8%. In addition, OG atom of Ser580 might form a salt bridge with NE1 atom of Trp485 (W485), possibly stabilizing the structure of the pore (Figure 3D). Substitution of the G580 amino acid with a Cysteine (G580C, Pt 14) introduces a bulkier residue at position 580 and changes its RSA from 5.9% to 3.7%. Although the free SH residue of C580 can make disulphide bonds with other amino acids with free SH residues, the distance between C580 and C486, which is the closer amino acid with a free SH residue (10.519Å; Figure 3E) does not allow the making of disulphide bonds. The R660T variant (Pt 15) replaces a buried charged Arginine (RSA 5.6%) with an uncharged Threonine. This substitution also disrupts a salt bridge between NH_2_ group of Arg660 and OD1 atom of Asp137 (distance: 4.787 Å) (Figure 3F). For V44M, R433H (Pt 2), V463I (Pt 11), and F697L (Pt 16), Missense3D suggests only mild alterations of the cavity volume (<70Å^3^), without significant structural changes, which might still influence the overall stability of the protein (Supplementary Table 3). It has indeed been demonstrated that cavities in membrane proteins play a pivotal role in balancing stability and flexibility, impacting protein function (15). For the T481N (Pt 3), which according to ConSurf affects a predicted structural residue, Missense3D did not indicate clear structural damage but could calculate cavity volume in the mutant structure. As for the in-frame deletion variant I475del (Pt 12), Phyre2 modelling did not predict gross alterations of the secondary structure.

### Post transcriptional editing in *TMEM63B* mRNA from human cerebral cortex and selection of variants for *in vitro* functional studies

We reverse-transcribed RNA from a human cerebral cortex sample into cDNA and amplified by PCR the region around exon 4 (Supplementary Figure 7A). We found that exon 4 was missing in 80% of *TMEM63B* RNA (Supplementary Figure 7B, C). We further characterised the Q/R editing at exon 20 in long and short *TMEM63B* isoforms by PCR and Sanger sequencing and found that the editing occurred only in the short isoform (Supplementary Figure 7D, E). Both the exon 4 splicing and Q/R editing findings are in line with previous observations in mice (10), except for lower Q/R editing occurrence in the human (∼40%) than in mouse cortex (∼80%) (Supplementary Figure 7E). Based on this, we decided to study the effects of selected *TMEM63B* variants *in vitro* by modelling them in the short non-edited isoform, which is the most abundant in the human cortex.

For our functional studies we selected three variants based on recurrence in multiple patients with overlapping clinical features (V44M) and on their location in critical transmembrane helices TM4 (R433H) and TM5 (T481N), whose rearrangement is implicated in channel opening (13).

### V44M, R433H and T481N do not affect TMEM63B localisation at the plasma membrane

The TMEM63B channel normally localises at the plasma membrane level (16, 17). To address the impact of V44M, R433H and T481N on TMEM63B localisation, we performed immunocytochemistry on Neuro2A cells transfected with GCaMP6f plasmids overexpressing either WT (wild-type) or mutant HA-tagged TMEM63B, and GCaMP6f empty vector as control.

We observed that, both in WT and mutant Neuro2A cells, TMEM63B was correctly localised at the membrane level (Figure 4). As expected, we did not observe any fluorescence emission in the red channel in GCaMP6f control cells. These findings suggest that all three variants we studied exert their effect by altering the channel function without impacting its localisation on the plasma membrane.

**Figure 4.**
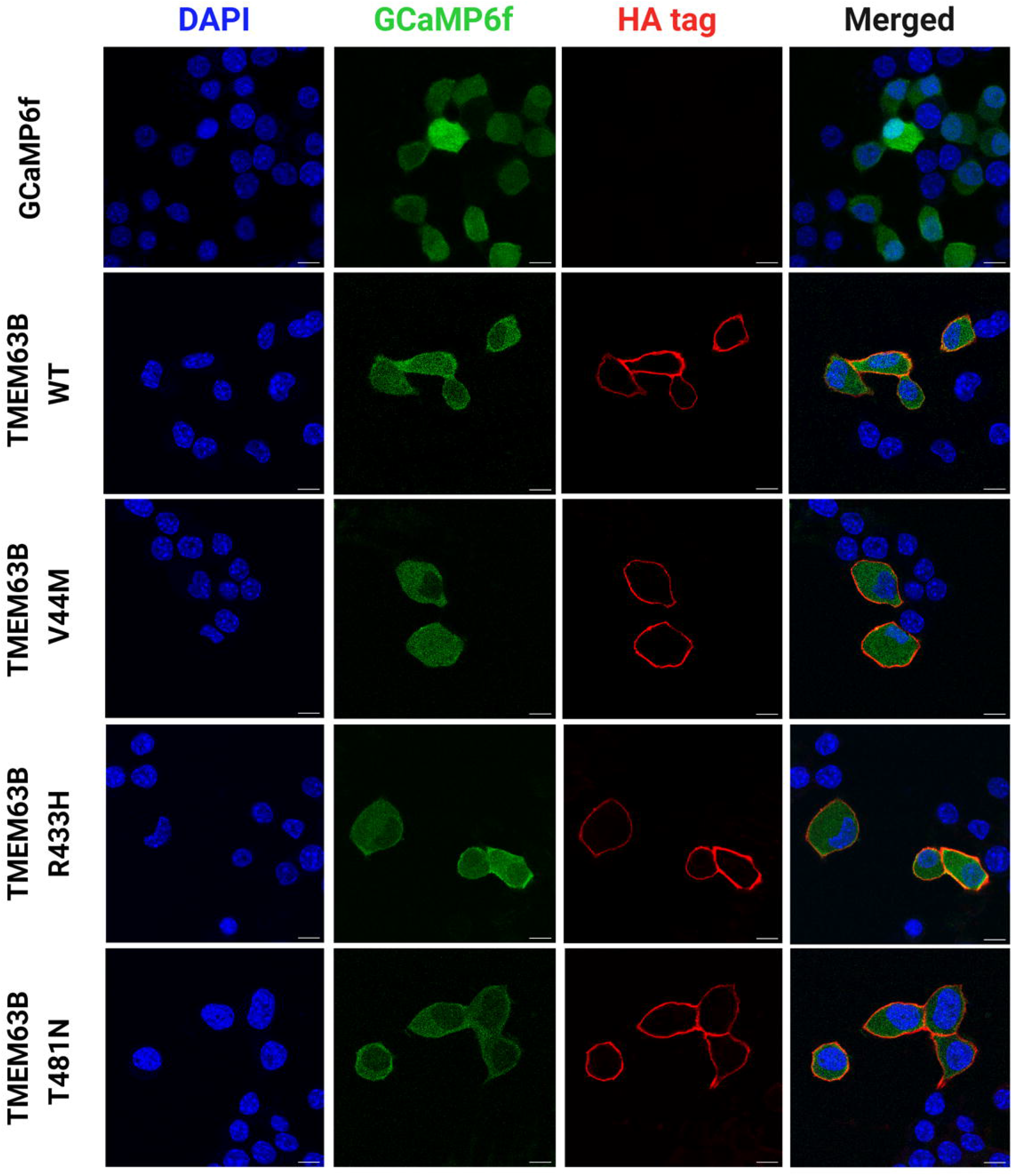
Immunocytochemistry to assess TMEM63B localisation at the plasma membrane. Confocal microscopy photographs of Neuro2A cells transfected with GCaMP6f empty vector, TMEM63B WT or mutant plasmids and analysed 48 hours post transfection. Transfected cells express the GCaMP6f protein which fluoresces in the green channel. Cells were stained with primary anti HA tag antibody, secondary Alexa Fluor 555 antibody and DAPI. Scale bar = 10 μm.

### V44M, R433H and T481N affect TMEM63B conductance

In physiological conditions, TMEM63B operates as a non-selective cationic channel activated by mechanical and osmotic stimuli (1, 4). To address the impact of the selected V44M, R433H, and T481N variants on channel properties, we applied a set of electrophysiological protocols previously implemented to characterise WT TMEM63B *in vitro* (4) (Figure 5, Supplementary Figure 8). In transfected Neuro2A cells, we initially recorded whole-cell currents elicited by a -80 to +80 mV voltage ramp protocol in isotonic conditions (300 mOsm/L extracellular solution) and then imposed a hypo-osmotic stimulus by switching to a 170 mOsm/L solution.

**Figure 5.**
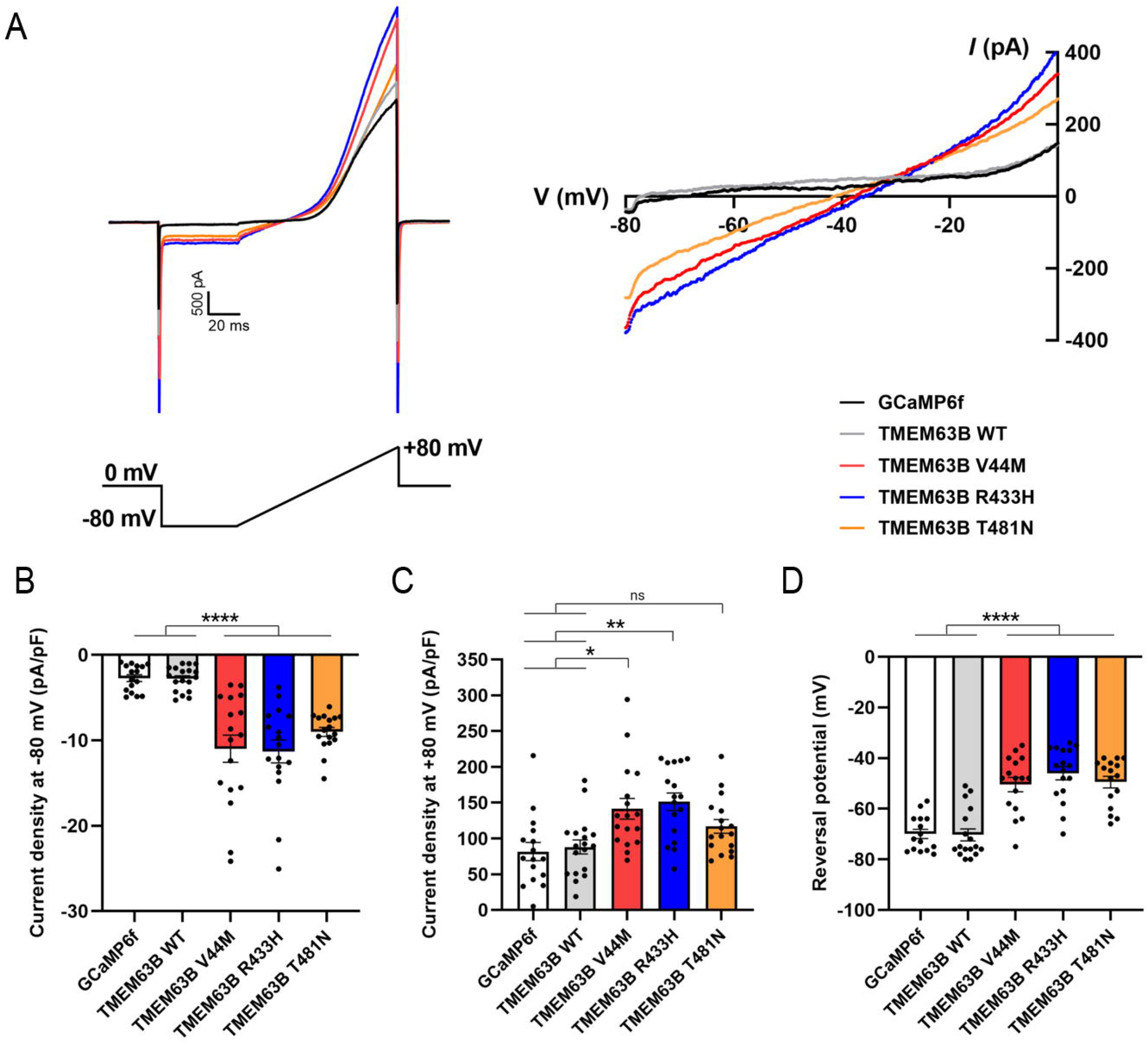
Electrophysiological recordings on transfected Neuro2A cells. (A) Left panel: representative raw traces of TMEM63B-mediated currents registered under isotonic condition in Neuro2A cells transfected with GCaMP6f, TMEM63B WT or mutant plasmids. Cells were held at 0 mV and recorded with a ramp protocol from −80 mV to +80 mV, 100 ms duration, 0.1 Hz. Right panel: current-voltage (I-V) relationship showing variant-induced change in reversal potential. (B-C) Quantification of whole-cell current density at -80mV (B) and +80mV (C) (GCaMP6f = 16 cells, WT TMEM63B = 18 cells, TMEM63B V44M, R433H and T481N = 17 cells; ****p<0.0001, **p<0.01, *p<0.05, ns = not significant, Kruskal–Wallis and Dunn’s multiple comparisons tests). (D) Quantification of reversal potential (GCaMP6f= 15 cells, WT TMEM63B = 17 cells, TMEM63B V44M = 16 cells, TMEM63B R433H = 17 cells, TMEM63B T481N = 16 cells; ****p<0.0001, One-way ANOVA and Tukey’s multiple comparison test).

In mutant TMEM63B cells, a -80 mV step imposed under isotonic conditions elicited a stable inward current that was absent in WT TMEM63B or GCAMP6f control cells (Figure 5A, B). Outward currents elicited at +80 mV were also significantly enhanced in cells expressing V44M, R433H, and T481N mutants compared to controls (Figure 5C). These three variants also caused a significant depolarization of the reversal potential, as shown by the current-voltage (I/V) relationship (Figure 5A, D).

Under hypo-osmotic conditions, a -80 mV step elicited an inward current in cells expressing either the WT or mutant TMEM63B, while no current was generated in control cells (Supplementary Figure 8). In most cases, cells underwent massive swelling and eventually burst, as previously reported (4). WT and mutant cells that sustained the hypo-osmotic shock and reached the plateau currents exhibited similar current amplitudes (Supplementary Figure 8A, B). The overall increase in currents observed at -80 mV, expressed as the delta (Δ) between currents measured under hypo-osmotic and isotonic conditions, was larger in WT than in mutant TMEM63B expressing cells (Supplementary Figure 8C), as might be expected by a leak inward current under isotonic conditions in the mutant.

Overall, these results show that V44M, R433H and T481N alter channel conductance, resulting in partial channel activation in physiological conditions, without altering the maximal channel conductance under hypo-osmotic stimulation.

### *TMEM63B* variants affect Calcium permeability

Previous experiments conducted on Neuro2A cells led to identify TMEM63B as an osmosensitive nonselective cation channel activated by hypo-osmotic stress (4). In cochlear hair cells, where TMEM63B is particularly enriched, the Ca^2+^ response is an essential element of the volume regulatory mechanism that protects cells viability from the osmotic challenge. Therefore, we wondered whether the clinically relevant *TMEM63B* variants might affect the hypo-osmotic induced Ca^2+^ responses.

Hypo-osmotic challenges trigger cationic currents across the TMEM63B channel, with hyposmolarity-induced Ca^2+^ influx (4). To determine whether the TMEM63B variants affect the hyposmolarity-induced Ca^2+^ response, we co-expressed in Neuro2A cells the Ca^2+^ sensor GCaMP6f and either WT TMEM63B or one of the three mutants V44M, R433H, and T481N. We performed calcium imaging under a confocal microscope in a recording chamber equipped with a perfusion system to deliver the test hypo-osmotic solutions to the cells. As previously reported (4), a hypo-osmotic stress provided by decreasing the osmolarity of the perfusion solution from 300 mOsm/L to 170 mOsm/L triggered a Ca^2+^ response (Figure 6A) in about 35% of WT TMEM63B cells, while only a small fraction of control cells transfected with GCaMP6f responded (Figure 6B). Neuro2A cells carrying V44M showed reduced responsivity with respect to the WT. We also tested the response to weaker osmotic stresses that are more likely to be representative of physiological changes, by shifting solution osmolarity from 300 mOsm/L down to 255 mOsm/L. The percentage of TMEM63B responsive cells decreased and all the three mutants showed a reduced responsivity score, which was significant for R433H (Figure 6B). The time course of representative responses plotted in Figure 6A,C shows a heterogeneous behaviour of cells, ranging from transient Ca^2+^ pulses to sustained increments.

**Figure 6.**
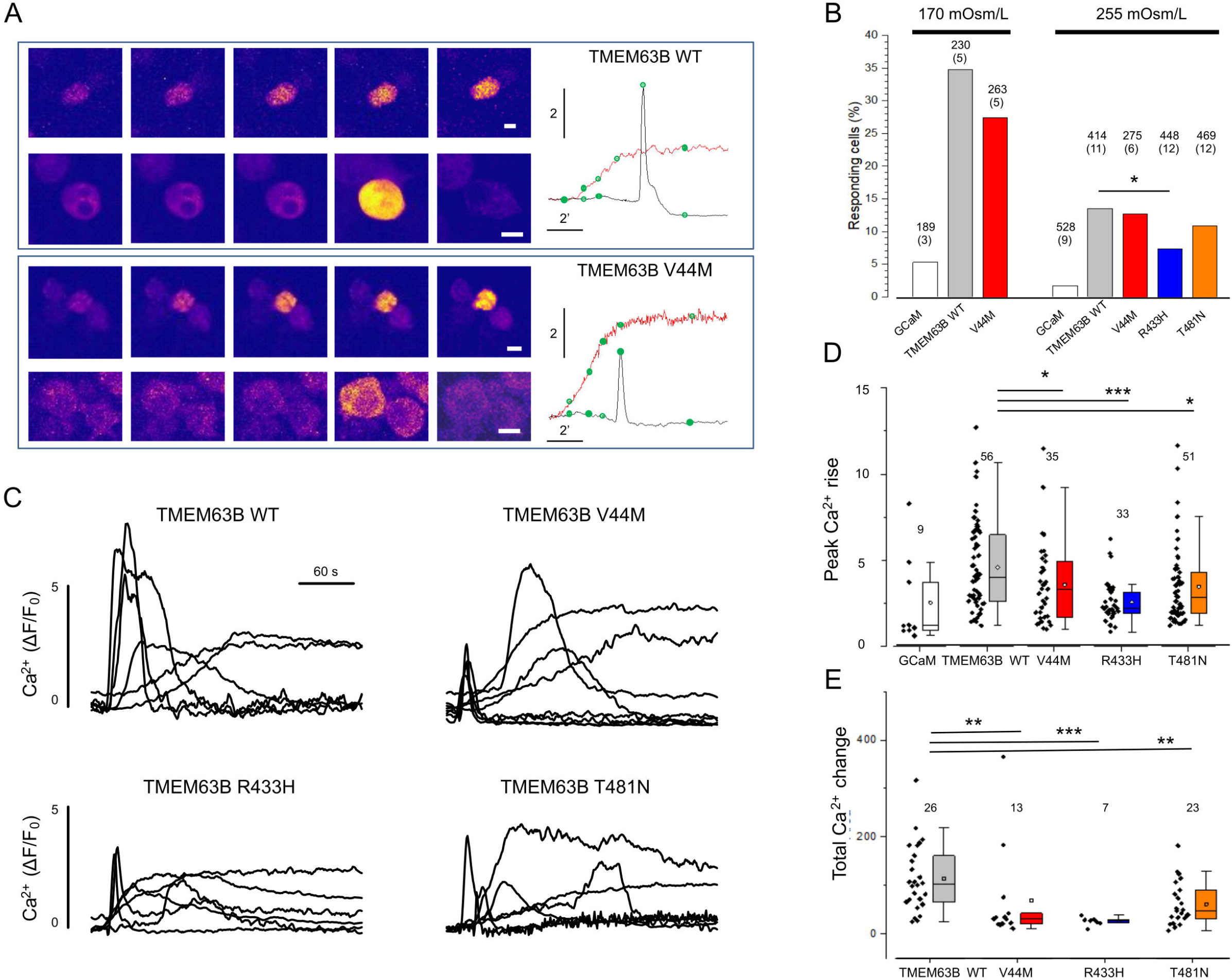
Mutations of TMEM63B impair the Ca^2+^ response to hypotonic stress. (A) Representative time-lapse sequence of Neuro2A cells co-transfected with WT and V44M TMEM63B after exposure to hypo-osmotic solution (170 mOsm/L). For each genotype we show two cells characterized by transient or steady responses. The green dots on the traces indicates the timing of the images. Calibration bar is 10 μm in all images. (B) Fraction of Neuro2A cells transfected with WT, V44M, R443H, or T481N TMEM63B presenting a Ca^2+^ response within 10 min from exposure to hypo-osmotic solutions. The number of analysed cells is on top of each column, with the number of replicates in parenthesis. (C) Representative traces showing the change of fluorescence for several cells transfected with WT, V44M, R443H, or T481N TMEM63B. (D) Cumulative results indicating the peak amplitude of the Ca^2+^ response to hypo-osmotic stimulus (255 mOsm/L). Numbers indicate the responding cells in each group. Total number of analysed cells and replicates as indicated in panel B. (E) Integral of the Ca^2+^ change for the indicated experimental groups. In this analysis we included only the cells that returned to baseline within 150 s from the transient onset. All mutant showed a drastically reduced response. Abbreviations and symbols: ***, p<0.001; **, p<0.01; *, p<0.05 (Chi-Square test); GCaM, control cells transfected with GCaMP6f empty vector.

Given the role of Ca^2+^ in orchestrating the cell response to osmotic stress (4), we next analysed the amplitude and dynamics of the observed Ca^2+^ changes. All three variants showed smaller Ca^2+^ transient compared to cells expressing WT TMEM63B. This is true when considering the maximal amplitude of the response (Figure 6D) and when considering the integral of the Ca^2+^ change computed for cells that returned to baseline within 150 seconds from the response onset (Figure 6E).

### V44M and G580C cause lethal toxicity in *Drosophila*

About two-thirds of the vital genes in the *Drosophila* genome are involved in eye development, including genes required for general cellular processes. For this reason, the fly eye provides an excellent experimental system to study the role of target genes in cellular function and development and in neurodevelopment/degeneration (18). To evaluate the potential impact of selected variants on the eye morphology, we generated transgenic flies expressing the human *TMEM63B* gene. We designed transgenes for the WT *TMEM63B*, the recurrent V44M, and the G580C variants, using the GMR-Gal4 ectopic expression system, which is mostly expressed in the retina (19). A schematic representation of the transgene construct is shown in Supplementary Figure 9A. The Gal4-UAS system (20) induces *TMEM63B* expression by the binding of the yeast transcription factor Gal4 to the UAS sequence. The expression level of the Gal4/UAS system increases in a temperature-dependent manner (19) and can therefore be modulated by changing the fly rearing temperatures. In WT *TMEM63B*-expressing flies, which were viable and reached the adult stage, we did not observe eye abnormalities (Supplementary Figure 9B, C). However, when expressing the V44M and G580C *TMEM63B* transgenes, we did not obtain any adult fly as both variants caused early lethality. We could not obtain any adult fly even after reducing the expression level of the two transgenes to the minimum by lowering the rearing temperature down to 18°C. In adult flies expressing WT *TMEM63B* by *c739-Gal4*, we performed immunohistochemistry, and observed dotted signals on the cell membrane (Supplementary Figure 9D), thus demonstrating correct expression of the transgene in a neuronal cell type, Kenyon cells. We observed the same lethal phenotype by expressing the *TMEM63B* transgenes under either the *GMR-Gal4* or the *c739-Gal4* drivers, demonstrating that both variants caused lethal toxicity in *Drosophila*. Since the lack of either Kenyon cells or photoreceptors is not sufficient in itself to cause death in flies, we expect this phenotype to be caused by the expression of the V44M and G580C *TMEM63B* transgenes in other cell types indispensable during developmental stages.

## Discussion

This series of 16 patients with pathogenic heterozygous *de novo* variants of *TMEM63B* defines the phenotypic features of an autosomal dominant early-onset DEE syndrome. All patients exhibited global developmental delay, moderate to profound intellectual disability, severe motor impairment, severe epilepsy with onset from birth to 3 years. Two patients had died prematurely and those who had reached adolescent or adult age remained completely dependent. Most had central visual impairment and swallowing dysfunction requiring PEG insertion. Epilepsy was a prominent feature and was quite severe at onset, featuring recurrent episodes of status in some patients, although a variable degree of seizure control was reported, with three patients achieving seizure-freedom on treatment. We noticed a common trajectory of the epilepsy phenotype, especially with the recurrent V44M variant, characterised by neonatal onset of apnoeic/focal seizures, evolving to epileptic spasms around the age of 4 to 6 months, and then continuing with focal, generalised seizures, or both. EEGs were consistent with an EE pattern at onset, including multifocal interictal epileptiform abnormalities and multiple seizure types in most. MRIs showed a consistent pattern of widespread or multiregional white matter abnormalities, with posterior periventricular predominance, dysmorphic lateral ventricles, thin corpus callosum, variably accompanied by global cerebellar atrophy (eight patients) and areas of cortical atrophy (six patients). Overall, initial MRI findings in most patients could be misdiagnosed as consequence of perinatal hypoxic-ischemic cerebral injury (see for example Pts 1, 3, 5, 7, 8, 12-15). However, longitudinal neuroimaging with relatively long-time interval (up to 13 years) often revealed signs of progression, especially affecting the white matter and cerebellum. Over a clinical monitoring period up to ages 20 months to 30 years, two patients died at ages 0-5 years and 11-15 years due to pneumonia. In the remaining 14 patients, clinical findings were consistent with a DEE akin to cerebral palsy in most, with limited signs of progression in ten (Pts 1, 2, 5, 7-9, 12, and 14-16), and of a milder encephalopathy in four (Pts 3, 10, 11, and 13).

Overall, clinical and imaging findings are consistent with a progressive neurodegenerative clinical course and indicate, in most patients, prenatal central nervous system impairment leading to onset of symptoms early after birth or within the first year of life. Haematological abnormalities might have contributed to brain damage. In 12 patients, we found macrocytosis or signs of chronic haemolysis, often from birth, with a fluctuating course (Table 1, Supplementary Table 1). Although such abnormalities were mostly not clinically significant or only caused jaundice in the neonatal period, in four patients the anaemia was so severe to require periodic transfusions. In four of the patients with anaemia, MRI showed progressive thickening of the trabecular (spongy) bone of the skull (Pts 4, 8, and 12 in Figure 1 and Patient 9, not shown), a finding often associated with severe haematological disorders (21).

*TMEM63B* is evolutionarily conserved and highly intolerant to both loss-of-function and missense variants in the general population. The International Mouse Phenotyping Consortium (IMPC, https://www.mousephenotype.org/) lists *Tmem63b* among the essential genes in database since its homozygous knockout causes pre-weaning lethality in the C57BL/6N strain (22). Heterozygous *Tmem63b* mutant mice from the IMPC exhibits a neurodevelopmental phenotype and increased circulating bilirubin level, both consistent with observations in our patients. No further significant haematological or sensorial abnormalities were observed in this mouse model, but susceptibility to epilepsy (e.g., by Electroconvulsive Threshold Testing phenotypic assays) was not assessed. A more vital C57BL/6N-FVB/N mixed breed surviving *Tmem63b*-knockout mice developed progressive hearing loss (4), a phenotype we did not observe in our patients.

Mendelian disease genes associated with autosomal dominant disorders are overrepresented among genes whose loss causes early-development lethality (DL) in mice (23). *TMEM63B* was prioritised among the potential candidates for human developmental disorders based on the overlap between genes that are DL in mouse and those carrying *de novo* variants in large-scale human rare disease sequencing data sets (23). Of five individuals with *de novo TMEM63B* variants included in the DDD (24) and 100KGP (25) studies, with minimal phenotypic information, we could retrieve detailed clinical data in three, which we added to this series (Pts 4, 6, and 7, Table 1).

All ten variants were distributed in a transmembrane domain conserved among osmosensitive calcium-permeable cation channels and affected residues under selective pressure in the general population, consistently with their predicted pathogenicity. None was a clear loss-of-function/haploinsufficient variant (e.g., causing frameshift or premature stop codon). Heterozygous deletions including *TMEM63B* are rarely reported in the Database of Genomic Variants, where at least two individuals carry truncating deletions removing most of the gene (http://dgv.tcag.ca/; supporting variants nssv1153700 nssv538990). Through our matchmaking initiative, we became aware of a patient with an unrelated phenotype and a homozygous truncating variant of *TMEM63B* (c.973C>T, R325*), present in both healthy, heterozygous parents (unpublished data). These observations argue against haploinsufficiency being the obvious pathogenic mechanism in our cohort, as also supported by structural modelling and functional data. Structural modelling suggested mild changes in 8/10 of the variants, without a disruptive effect on the protein structure. All the ten variants affected transmembrane helices, including TM4-TM8, constituting the channel pore, or TM1 and TM7, involved in sensing membrane tension (2), and were predicted to cause minimal structural changes, such as disruption or creation of single salt bridges, and changes in cavities volumes. The predicted mutational consequences might therefore imply altered channel function, e.g. by stabilizing the pore opening or affecting ion permeability and selectivity. In line with this hypothesis, and as expected for variants sparing protein folding, none of the three variants we tested *in vitro* impaired protein localisation in the plasma membrane.

V44M was recurrent in seven patients and the remaining variants clustered across a ∼270 residues transmembrane region. Recurring mutations, as well as mutations with spatial clustering patterns, may exert their pathogenic effect through disease mechanisms other than LoF, such as gain-of-function (GoF) with enhanced activity or dominant-negative effects (26, 27).

In two independent *Drosophila* models, loss of *Tmem63* resulted in viable flies, not exhibiting gross defects in coordination, but lacking the ability to discriminate food texture or humidity (28, 29). In one of these models, the ectopic expression of the human *TMEM63B* gene in knockout flies rescued the defective phenotype in moisture attraction, demonstrating functional conservation of the two orthologues (29). We thus decided to model the V44M and G580C variants in *Drosophila*. However, both V44M and G580C TMEM63B transgenes caused lethal toxicity in our models. In contrast, the phenotype of the WT *TMEM63B-*expressing flies was indistinguishable from the control animals. These observations suggest that the variants we tested cause a toxic GoF of TMEM63B, rather than LoF.

Heterozygous variants of *TMEM63A* have been associated with developmental delay and hypomyelination (6–8). Disease-associated missense variants are enriched at amino acid sites that are conserved across paralogues (30) and differences in the clinical consequences of variants in paralogues may be due to different expression patterns and novel functions emerged with adaptive evolution (31). When aligning protein sequence of, and comparing variants positions in, *TMEM63B* and *TMEM63A* (Figure 1A, Supplementary Figure 2), we found that the Glycine 580 mutated to Serine in Pt 11 and to Cysteine in Pt 13 corresponds to the Glycine 567 recurrently mutated to Serine in *TMEM63A* in two unrelated patients with transient hypomyelination (6). G580S affects a buried Glycine in TM7. In the Arabidopsis AtOSCA1.1 channel the corresponding Glycine 528 is located in a bending of the TM6 helix (2). Targeted mutagenesis of Glycine 528 to Alanine or Proline reduces the pressure necessary to elicit a current, suggesting that channel activation might involve straightening of M6 around Glycine 528 to relieve the blockage of the ion channel pore (2). According to *in silico* prediction and structural modelling, we might expect that both variants of Glycine 580 stabilize the structure of the pore.

Multiple expression datasets indicate a complementary expression pattern between *TMEM63A* and *TMEM63B*, with the first being mainly expressed in oligodendrocytes and the second strongly expressed in neurons and to a lesser extent in astrocytes and oligodendrocyte precursor cells (OPC) (Supplementary Figure 10; https://singlecell.broadinstitute.org; https://www.proteinatlas.org/). The six patients reported to date with *TMEM63A* variants had follow-ups of variable duration and exhibited phenotypes that were either similar to our patients’ (7, 8) or milder in those whose white matter changes improved with age (6), a phenomenon we did not observe.

For *in vitro* functional studies, we focused on the *TMEM63B* mRNA isoform most represented in the human cerebral cortex, the main generator of epileptogenic activity. Previous experiments in transfected Neuro2A cells suggested that the WT TMEM63B channel mediates non-selective cationic currents in response to hypo-osmotic stimulation (4). For the *in vitro* study of V44M, R433H and T481N, in line with to previous approaches (4) we used Neuro2A cells and adopted an electrophysiological protocol consisting of a voltage ramp from -80 mV to +80 mV. In physiologic isotonic conditions, the -80 mV step revealed leak inward currents in cells expressing mutants but not WT TMEM63B or control GCaMP6f, indicating that the variants lead to a gain in conductance even in the absence of the hypo-osmotic stimulus gating the channel.

Under a hypo-osmotic challenge, we measured similar values of inward current amplitude in both mutant and WT *TMEM63B* expressing Neuro2A cells, suggesting that the variants do not alter the maximal channel conductance. However, the overall increase between currents measured in hypo-osmotic and isotonic conditions was larger in WT than in mutant *TMEM63B* expressing cells. Similarly, the Ca^2+^ transients generated under hypo-osmotic stimulation were smaller across the mutant channels compared to the WT. One possible explanation for these observations is that V44M, R433H and T481N result in a selective reduction of the relative permeability for Ca^2+^ in favour of Na^+^.

Inward cationic leak currents in neural cells expressing TMEM63B mutants may lead to altered neuronal excitability and/or impaired Ca^2+^ homeostasis. In C57BL/6N-FVB/N *Tmem63b* knockout mice, progressive hearing loss due to necroptosis of the outer hair cells (OHCs) (4), which face severe shape- and volume-changing conditions, was suggested to reflect an abnormal response to osmotic and mechanical stimuli, ultimately leading to cell death (4).

Brain shrinking with white matter changes, the main imaging finding in our series, indicates defective myelination and, possibly, defective oligodendrocytes development. Clinical and imaging findings suggest that cell damage is already present at birth but further progresses postnatally. Already during prenatal life, several factors may determine osmotic challenges to the brain, including hydration changes, electrolytes imbalance and mechanical stress. In a brain where neural cells are exceedingly vulnerable to even trivial volume changes and electrolytic imbalances, recurrent seizures may, in a vicious circle, make *TMEM63B-*defective neurons particularly susceptible to osmotic imbalance. During seizure activity, the extracellular environment surrounding axons and oligodendrocytes undergoes changes in volume and osmolarity (32) and, given the very small volume of the extracellular space surrounding myelinated axons, oligodendrocytes might be subjected to continuous changes in local osmolarity. Unfortunately, our understanding of the homeostasis of the extracellular space is still in its infancy (33). In line with the above considerations, the epilepsy phenotype observed in our cohort is considerably more severe compared to other neurological conditions featuring structural damage of similar magnitude (i.e., cerebral palsy due to vascular injury) (34).

In 12 patients, we also observed haematological abnormalities of variable severity, with signs of chronic haemolysis and myelodysplasia. In inherited haemolytic anaemias, abnormally shaped RBCs may reflect disorders of cation permeability in the membrane, resulting in cellular over- or de-hydration. Dehydrated hereditary stomatocytosis (DHSt), a rare congenital haemolytic anaemia, is caused by dominant GoF mutations of *PIEZO1*, encoding for a stretch-activated ion channel (35, 36). In some patients with DHSt, hematologic abnormalities are subtle (35, 36), as also seen in most of our patients in whom they were revealed. In DHSt, *PIEZO1* mutations keep the channel in an open conformation with prolonged activity, resulting in Ca^2+^ influx and consequent K^+^ efflux though Ca-sensitive Gardos channels, decreasing intracellular osmolarity and causing dehydration of RBCs (36, 37). Although the role of *TMEM63B* in RBCs remains to be elucidated, it is known to be expressed in the RBC membrane (http://rbcc.hegelab.org/) like *PIEZO1* and might act similarly.

Erythrocytes are highly deformable cells. The intense mechanical solicitations they face when circulating through the brain microvasculature require highly effective mechanosensory feedback mechanisms. Under normal healthy conditions, deformability of their membrane allows RBCs to flow through vessels of diameter less than the cell diameter (7-8 μm), ensuring robust tissue perfusion and oxygen delivery (38). This RBCs feature is particularly critical in the brain microvascular network, where the diameter of cortical capillaries is around 4-5 μm (39). RBCs also play an active role in stabilizing neurovascular flow dynamics, both by their physical properties (40) and by targeting the vascular endothelium with vasoactive molecules (41). Chronic anaemic changes and the physical vulnerability of *TMEM63B-*defective RBCs might contribute to white matter abnormalities, which often predominate in watershed vascular areas in the patients reported here (Figure 1). Similar cerebrovascular complications are known to occur in patients with hereditary haemolytic anaemia (42).

In conclusion, TMEM63B developmental and epileptic encephalopathy represents a novel, clinicopathological entity deriving from the dysfunctional behaviour of a highly conserved stretch-activated ion channel. *TMEM63B* variants cause a gain in conductance with impaired Ca^2+^ homeostasis and are associated with neurodegenerative changes that start during prenatal brain development and slowly progress during postnatal development and adulthood, affecting the cerebral white and grey matter, and cerebellum. The clinical counterpart of such widespread anatomic abnormalities includes severe early onset epilepsy, associated with moderate to profound intellectual disability and severe motor and cortical visual impairment. Although a clear genotype-phenotype correlation is not yet possible, the phenotype associated with the recurrent V44M variant is uniformly severe. Concomitant haematological changes make this a complex syndrome also featuring potentially life-threatening acute haemolytic episodes occurring without obvious triggers.

## Methods

### Patients

Our initial discovery cohort consisted of 600 consecutive patients referred to the Neuroscience Department of the Meyer Children’s Hospital to investigate the genetic causes of DEEs. Within this cohort, we identified by whole exome sequencing (WES) (43) *de novo* heterozygous variants of *TMEM63B* in three patients (Pts 1-3) with a homogenous clinical and neuroimaging phenotype and promoted an international collaboration through GeneMatcher (44) identifying 13 additional patients carrying *de novo TMEM63B* variants (Pts 4-16). We reviewed medical records, EEGs, and brain MRI scans. We classified seizure types following the ILAE criteria (45), whenever applicable, and used more descriptive terms when seizure phenomenology could not fit classification terminology.

Detailed methods for genetic investigations, protein structural analysis, functional characterization of *TMEM63B* variants, and *Drosophila* modelling are reported in the Supplementary material.

### Statistical analysis

We used the GraphPad 8.0 software for statistical analyses. We assessed the normal distribution of experimental data using the D’Agostino-Pearson normality test. We analysed data with normal distribution with One-way ANOVA followed by Tukey’s multiple comparison test, while for non-normally distributed data we used the unpaired Mann-Whitney U test or Kruskal-Wallis test followed by Dunn’s multiple comparison test. We set significance level at p<0.05 and expressed data as mean ± standard error of the mean (SEM).

For statistical analysis of calcium imaging, we used the Origin 2019b package (OriginLab, Massachusetts, USA). Hypotheses were tested with Mann-Whitney non-parametric tests. We considered results to be significant for p<0.05.

### Study approval

We obtained written informed consent for the study from all participants or their legal guardians, according to local requirements. The study was approved by the Paediatric Ethics Committees of the Tuscany Region, Italy, in the context of the DESIRE FP7 EU project and its extension by the DECODE-EE project.

## Data Availability

Data availability
The data supporting the findings of this study are available within the article and/or its Supplementary material. Any additional raw data are available on request from
the corresponding author.

## Author contributions

RG designed the research study, acquired and analysed clinical and genetic data, and drafted the manuscript; AV participated in designing research study, analysed genetic data, and drafted the manuscript; SB acquired and analysed clinical data, and participated in drafting the manuscript; CP analysed functional data, performed statistical analysis, and participated in drafting the manuscript; GMR participated in designing functional studies in transfected cells, analysed calcium imaging data, and performed statistical analysis; VC participated in designing functional studies, and in genetic and protein structural data analysis; AM, GM, RP, and SG performed experimental analyses in transfected cells; KH and HO participated in protein structural data analysis; AD, CvS, EV, GB, JvdS, MK, MPM, MS, NM, RB, RR, SH, TB, and TR acquired clinical and genetic data; EA, EKB, JT, KW, MSc, RK, and SV acquired clinical data; AT, DM, DS, EHS, KM, KvG, MJVH, PC, and TC acquired genetic data; AS and TS produced and analysed *Drosophila* models.

## Data availability

The data supporting the findings of this study are available within the article and/or its Supplementary material. Any additional raw data are available on request from the corresponding author.

## Acknowledgments

We are grateful to all the Patients and family members for their participation in this study. This work was generated within the European Reference Networks EpiCARE and ITHACA.

