## Supplementary for "Stretch-activated ion channel TMEM63B associates with developmental and epileptic encephalopathies and progressive neurodegeneration"

### Sections:

### Supplementary Materials and Methods

#### Genetic investigations

We performed whole exome (WES, Pts 1-3, 5, and 7-16) and genome (WGS, Pts 4 and 6) sequencing using standard procedures on DNA extracted from peripheral blood. In all but Pts 7 and 10, whose DNA was sequenced as singleton, we used a patient-parent trio sequencing strategy. We prepared DNA libraries by different kits according to manufacturers' instructions and performed paired-end sequencing on Illumina sequencers (Illumina, San Diego, CA, USA). We aligned sequencing reads to the human reference genome build GRCh37/hg19 by the Burrows-Wheeler Alignment (BWA-MEM) software package (1) and followed the Genome Analysis Toolkit Best Practices workflow (2) for variant calling. Detailed sequencing and variant annotation methods for each patient are provided as a reference to previous publications in the Supplementary Table 4. For variant analysis, we focused on exonic/splice-site single-nucleotide variants (SNVs) and coding insertions/deletions (InDels) with minor allele frequency (MAF) lower than 0.01 in the GnomAD v2.1 (http://gnomad.broadinstitute.org/) (3) or TOPMed (https://bravo.sph.umich.edu/freeze3a/hg19/) datasets. We excluded population-specific variants by interrogating our internal database (singleton WES data from approximately 2000 patients with DEE). We evaluated the potential impact of SNVs and InDels by the pre-computed genomic variants score from dbNSFP (4) and by the evolutionary conservation scores (5, 6). After filtering and interpretation, we proceeded to validation of the *TMEM63B* variants by Sanger sequencing (primers and conditions available on request). We followed the nomenclature guidelines of the Human Genome Variation Society (HGVS, http://www.hgvs.org/mutnomen) and referred to the NM_018426.3 reference transcript.

We evaluated *TMEM63B* gene-level constraint scores according to GnomAD (3) and the region-level constraint scores according to the Metadome and MTR tools (7, 8).

#### Homology modelling and structural analysis

To evaluate the evolutionary conservation of the mutated residues, we obtained the protein sequence of human TMEM63B, its paralogues TMEM63A and TMEM63C, and orthologues in five different vertebrate species (*Pan troglodytes, Sus scrofa, Mus musculus, Gallus gallus, Danio rerio*) from the NCBI Protein Database (9), and aligned them using Clustal Omega (10). As the protein crystal structure of TMEM63B has not been determined, we used the protein homology recognition engine Phyre2 (11) to predict and analyse the protein structure. For structural modelling, we referred to the NP_060896.1 human TMEM63B protein sequence using the Phyre2 'intensive mode' prediction option. To further analyse possible effects of the recurrent V44M, we calculated the free energy change due to this substitution by the FoldX software (12, 13) using a TMEM63B structural model predicted by AlphaFold2 (14). We also generated a tridimensional model incorporating evolutionary sequence conservation with the ConSurf webserver (15) with default parameters. We used the Missense3D tool (16) to predict possible structural changes introduced by the missense substitutions. To graphically represent the variants identified in our patients on the homology-predicted protein model we used the UCSF Chimera Visualization System (17).

#### RNA reverse transcription and cDNA analyses

We reverse-transcribed total RNA from healthy adult human cerebral cortex (BioChain, Newark, USA) into cDNA using the High-Capacity RNA-to-cDNA Kit (Applied Biosystems, Waltham, USA). We performed polymerase chain reaction (PCR) on the cDNA using the FastStart Taq DNA Polymerase (Roche, Basel, Switzerland) and *TMEM63B* primers designed with the Primer3 Plus software (https://www.bioinformatics.nl/cgi-bin/primer3plus/primer3plus.cgi) using the NM_018426.3 transcript as template. Primers and RT-PCR conditions are available upon request.

To characterise the alternative splicing of exon 4 in the *TMEM63B*, we amplified the cDNA region spanning the exon 4 (exons 3-8) and analysed the PCR product by agarose gel electrophoresis. We acquired the gel images by ChemiDoc Imaging System (Bio-Rad, Hercules, USA) and quantified the two cDNA bands we resolved by ImageJ software (National Institutes of Health, USA). We quantified the expression level of the two isoforms as the ratio of integrated densities of the two bands to the total. We extracted the cDNAs from excised gel bands (Macherey-Nagel, Düren, Germany), sequenced them and analysed the electropherograms using the SnapGene software (https://www.snapgene.com/) to confirm that they corresponded to the two *TMEM63B* isoforms with (herein referred to as “long” isoform) or without (herein referred to as “short” isoform) exon 4. As an orthogonal method to confirm the relative expression level of both isoforms, we also cloned the PCR product in TOPO TA cloning (Invitrogen, Waltham, USA) and used Sanger Sequencing to determine the number of colonies containing the long or the short isoform.

To characterise the Q/R editing at exon 20 in the *TMEM63B*, we amplified the cDNA region including exon 20 in both isoforms. To quantify the editing occurrence in both isoforms, we cloned the PCR products in TOPO TA cloning and counted the number of colonies containing the editing.

##### *TMEM63B* constructs

We designed wild type (WT) and mutant (V44M, R433H, and T481N) human *TMEM63B* cDNAs corresponding to the most represented isoform in the human cerebral cortex (short not edited), with the hemagglutinin (HA) tag sequence (AGCGTAATCTGGAACATCGTATGGGTA) at the 5' end. We obtained *TMEM63B* cDNAs, cloned into the pGP-CMV-GCaMP6f vector (Plasmid #40755, Addgene, Watertown, USA) (18), so that the cDNA is fused to the N-terminus of GCaMP6f via a P2A linker forming a tandem expression system (TMEM63B-P2A-GCaMP6f) (Genescript, Piscataway, USA). The correct inserts orientation and sequence validation was performed via Sanger sequencing by Genescript.

##### Cell culture and transfection

We maintained Neuro2A mouse neuroblastoma cells at 37°C in a humidified 5% $CO_2$ incubator in Dulbecco's Modified Eagle Medium (Invitrogen, Waltham, USA) supplemented with 10% foetal bovine serum (FBS) and 2 mM L-glutamine. For the electrophysiology experiments, we plated the cells onto 13 mm square glass poly-L-lysine coated coverslips. The cells used in the Ca-imaging experiments were plated on WillCo Wells dishes HBST-3512 (WillCo Wells B.V., Amsterdam, The Netherlands). We transfected Neuro2A cells using Lipofectamine 2000 (Invitrogen) according to manufacturer's instruction.

##### Immunocytochemistry and confocal microscopy

Forty-eight hours post transfection, we fixed Neuro2A cells in 4% paraformaldehyde in phosphate-buffered saline (PBS) for 15 minutes at room temperature, blocked with 10% normal goat serum and 0.1% bovine serum albumin (BSA) in PBS for one hour at room temperature and incubated in 0.1% BSA in PBS overnight at 4°C with the primary anti-HA tag antibody (1:500, #2367, Cell Signaling Technology, Inc., Danvers, USA). After washing in PBS, we incubated the cells in 0.1% BSA in PBS for one hour at room temperature with the secondary Alexa Fluor 555 antibody (1:500, #A21424, Thermo Fisher Scientific, Waltham, USA). We washed the cells in PBS and mounted coverslips with ProLong Gold Antifade Mountant with DAPI (#P36935, Thermo-Fisher Scientific). We acquired images of 100 x 100 µm area using a laser scanning confocal microscope (SP5, Leica, Wetzlar, Germany) and analysed data with ImageJ software (National Institutes of Health, Bethesda, USA).

In *Drosophila*, we performed immunohistochemistry and sample preparation as described previously (19). Briefly, we dissected the adult fly brains in PBS, fixed them in 4% formaldehyde (Electron Microscopy Sciences, USA), and incubated the samples with mouse anti-myc (4A6; 1:5000; Merck KGaA, Darmstadt, Germany) and secondary anti-mouse Alexa Fluor 568 (1:400; Thermo Fisher Scientific). We acquired images using a FV3000 confocal microscope (Olympus, Tokyo, Japan), and processed and analysed them using IMARIS 9.6.0 (Bitplane, Zurich, Switzerland).

#### Electrophysiology

Forty-eight hours post transfection, we recorded Neuro2A cells by whole-cell patch clamp. Recording pipettes were obtained from borosilicate capillaries (Harvard Apparatus, Holliston, USA) with a Narishige vertical puller (Narishige, Tokyo, Japan) and back-filled with a solution containing (in mM) 80 K-gluconate, 10 HEPES, 130 Mannitol (pH 7.4 with KOH, 300 mOsm/L), resulting in a bath resistance of 8-10 MΩ. We initially perfused cells with an isotonic extracellular solution containing (in mM): 80 Na-gluconate, 1 Ca-gluconate, 10 HEPES, 130 Mannitol (pH 7.4 with NaOH, 300 mOsm/L) at room temperature and with a flow rate of approximately 1 ml/min. The hypo-osmotic extracellular solution contained (in mM): 80 Na-gluconate, 1 Ca-gluconate and 10 HEPES (pH 7.4 with NaOH, 170 mOsm/L). The osmolarity of the solutions used in both patch clamp and calcium imaging experiments was measured by an osmometer (OSMOMAT 030, Gonotec Gmbh, Berlin, Germany). Solutions were exchanged with an eight-line valve switcher (Hamilton Company, Reno, USA). We stimulated the cells with a voltage clamp protocol consisting of a 40 ms negative step to -80 mV, followed by a 100 ms ramp stimulus to +80 mV, imposed every 10 seconds. Holding potential was 0 mV. Signals were sampled at 10 kHz and low pass filtered at 3 kHz with an Axon Multiclamp 700B

(Molecular Devices, Sunnyvale, USA). We performed data analysis offline with Clampfit 10 (Molecular Devices) and GraphPad 8.0 software (GraphPad, San Diego, USA).

#### Calcium imaging

Forty-eight hours post transfection, we acquired cells imaging with a ZEISS LSM800 Airyscan inverted microscope (ZEISS, Jena, Germany) at a resolution of 512x512 pixels (objective ZEISS ECPlan-NEOFLUAR 20x) with the standard settings for excitation and acquisition of Fluorescein (excitation 488nm, emission >500nm) at 0.5 Hz frequency for 600 seconds. During imaging, cells were exposed to solutions of variable osmolarity by perfusing the imaging chamber with peristaltic pumps (flux 1ml/min). The hypo-osmotic solution contained (mM): 65 NaCl, 5 KCl, 1 $CaCl_2$ 2, 1 $MgCl_2$ 2, 10 HEPES, pH=7.4, adjusted with NaOH. We prepared the isotonic solution adding mannitol to the hypo-osmotic solution to a final concentration of mannitol 130 mM without changing other ion concentrations.

#### Analysis of calcium imaging data

We analysed the acquired images with Fiji ImageJ software. For each field of acquisition, we generated an average projection, and we selected the cells which florescence was in the upper 95 percentile to exclude autofluorescent cells. We then analysed the selected cells by computing the fractional change of fluorescence defined as:

$$\Delta F(t) = \frac{F(t) - F_0}{F_0}$$

where $F_0$ indicates the baseline fluorescence computed as the average of the first 20 frames of the sequence. Cells were considered responsive when they presented changes of fluorescence larger than 50% of the baseline level.

#### *Drosophila* strains and generation of the transgenic lines expressing human *TMEM63B* WT, V44M, and G580C

Flies were maintained at 25°C on standard fly food. *GMR-Gal4* (#1104) and *c739-Gal4* with *UAS-CD8GFP* (#64305) were obtained from the BDRC (Bloomington, Indiana, USA). *40D-UAS* (#60101) for control experiments was purchased from VDRC (Vienna, Austria). To express human WT TMEM63B, V44M, and G580C, tagged with Myc at C terminal of the protein, the cDNAs were inserted into pJFRC81-10XUAS-IVS-Syn21-GFP-p10 (ID36432, Addgene, Massachusetts, USA) (Vectorbuilder Inc., Yokohama, Japan). We used the phiC31 integrase system (20) to insert the transgenes in the same position of the fly genome and exclude potential positional effects on gene expression. These vectors were inserted into the *aTTP40* landing site (WellGenetics, Taipei, Taiwan). As the expression level of the Gal4/UAS system increases in a temperature-dependent manner (21), the transgenic flies were reared at different temperatures (18°, 20°, 25° and 29°C), to modulate the expression of the transgene.

#### Eye imaging with bright-field microscopy and the quantification of morphological defects in the retina

WT *TMEM63B* and each the two variants were expressed by the *GMR-Gal4* driver and reared at 20℃. One-day-old flies were immobilized by freezing at -80℃ for imaging. These flies were imaged using a BX53 microscope system with a MPLFLN 20x objective lens (Olympus, Tokyo, Japan). The phenotypic scores were calculated using Flynotyper (22). Experimental analyses were performed using Prism 9 (GraphPad Software Inc., San Diego, CA, USA). The distribution of our data was determined using the D'Agostino & Pearson test and the Kolmogorov-Smirnov test (normality test was passed if $p > 0.05$). For data following a Gaussian distribution, we used ordinary one-way ANOVA with Tukey's multiple comparisons between groups.

#### Supplementary Figures

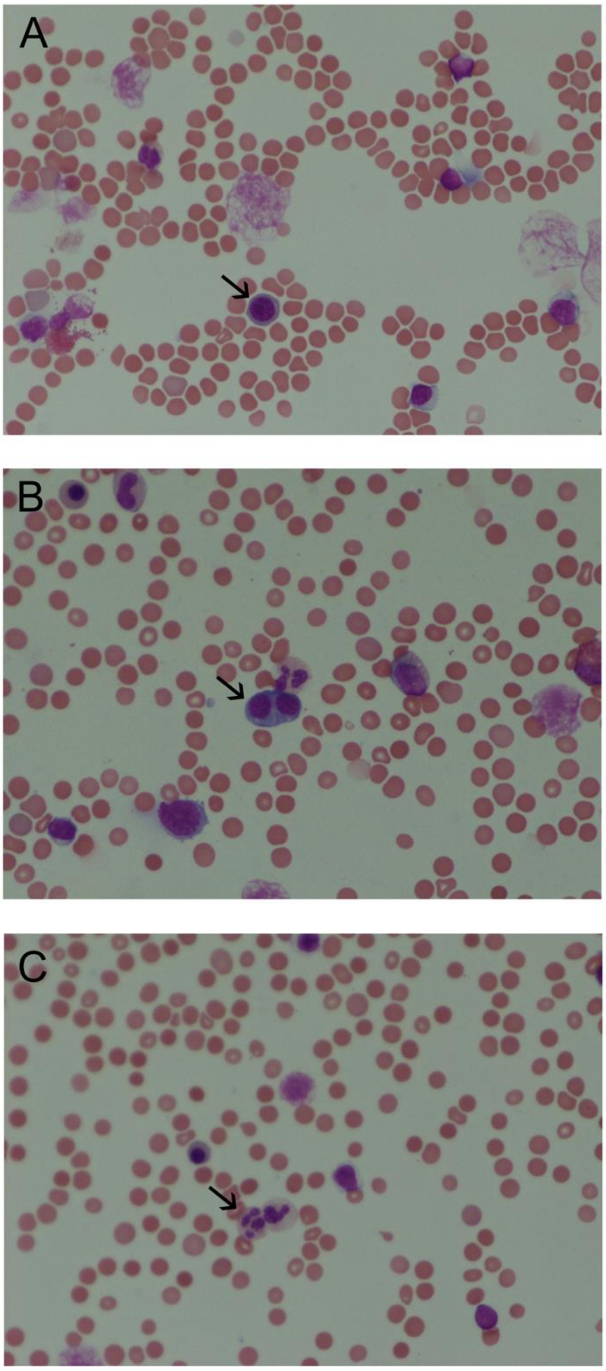

**Supplementary Figure 1 – Bone marrow aspirate smear from Patient 8**

Bone-marrow examination in Patient 8 (age 6-10y) revealed hypocellular marrow, signs of haemolysis, and evidence for haematopoiesis and myelodysplastic syndrome with aplastic anaemia. Arrows highlight examples of megaloblastic changes (A), binucleated erythroblasts (B), and Pseudo-Pelger–Huët anomalies (C) observed in the sample.

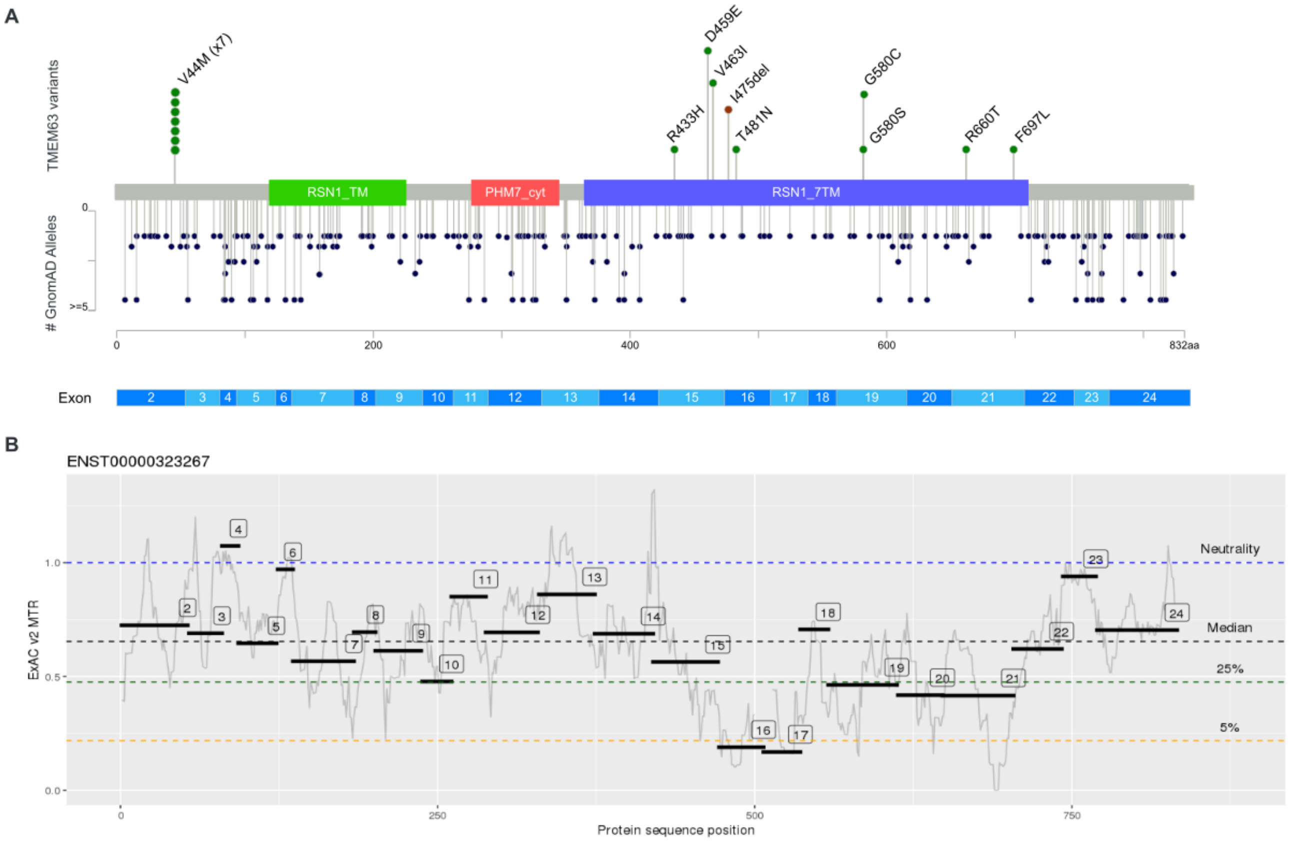

#### Supplementary Figure 2 - Distribution of *TMEM63B* variants in our cohort and in reference population

(A) The lollipop diagram shows the distribution of the *TMEM63B* variants observed in our cohort (top, missense variants in green, in-frame deletion in brown) versus the reference population in GnomAD 2.1 (bottom, missense variants all in dark blue) on the linear protein map and relative to the Pfam-identified domains (RSN1_7TM, PF02714, green; PHM7_cyt, PF14703, red; RSN1_TM, PF13967, blue) of the protein and the *TMEM63B* exons (NM_018426.3, ENST00000323267). All but one of the *TMEM63B* variants in our cohort map in the RSN1_7TM domain (PF02714), which is conserved among osmosensitive calcium-permeable cation channels (23). In the bottom panel, the length of the lollipop reflects the number of alleles in GnomAD 2.1 (see the Y-Axis on the left). The variants in our cohort maps in positions which are under constraint for missense variants. (B) The missense tolerance ratio (MTR)-Viewer tool (8) shows the local constraint with respect to the *TMEM63B* exons: all the variants in our cohort fall in regions where the MTR scores were below the neutrality threshold. The exon 16, bearing the T481N and the I475del variants, has the highest intolerance to missense substitutions, followed by the exons 15 (R433H, D459E, and V463I), 19 (G580S and G580C), 21 (R660T and F697L), and 2 (V44M).

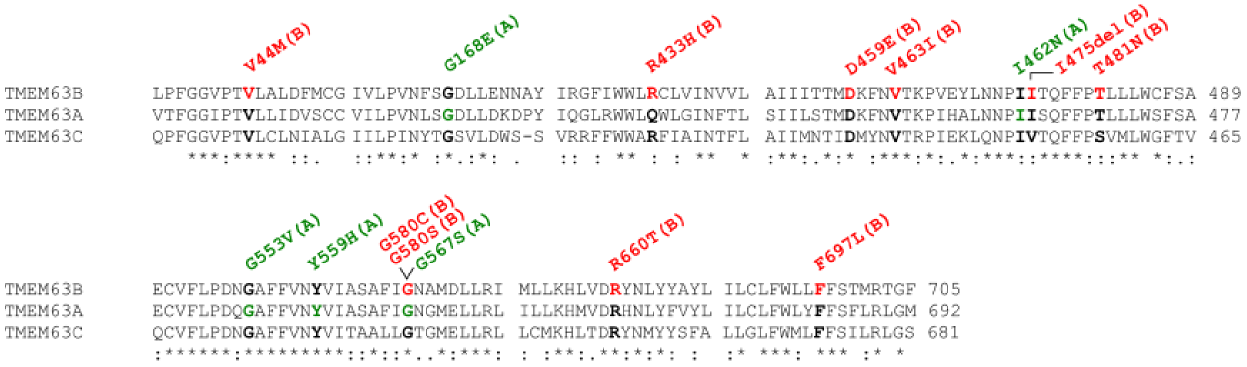

**Supplementary Figure 3 - Multiple sequence alignment of TMEM63A, B and C**

The alignment shows the protein sequence of the human TMEM63B protein (NP_060896.1) and of its two paralogues TMEM63A (NP_055513.2) and TMEM63C (NP_065164.2). The residues affected by heterozygous variants of *TMEM63B* (our cohort, red) or *TMEM63A* (from the literature, green) (24–26) are in bold. The details of the variants are displayed above the alignments. The asterisk below the tracks indicates positions which have a single, fully conserved residue between all the input sequences, the colon indicates conservation between groups of strongly similar properties, and the period indicates conservation between groups of weakly similar properties. Six out ten *TMEM63B* variants in our cohort affected residues which are fully conserved among all the three paralogues' sequences (V44, D459, V463, G580, R660, and F697), and three affected residues conserved among two of the three paralogues and maintaining strongly similar properties in the other one (R443, T481 and I475).

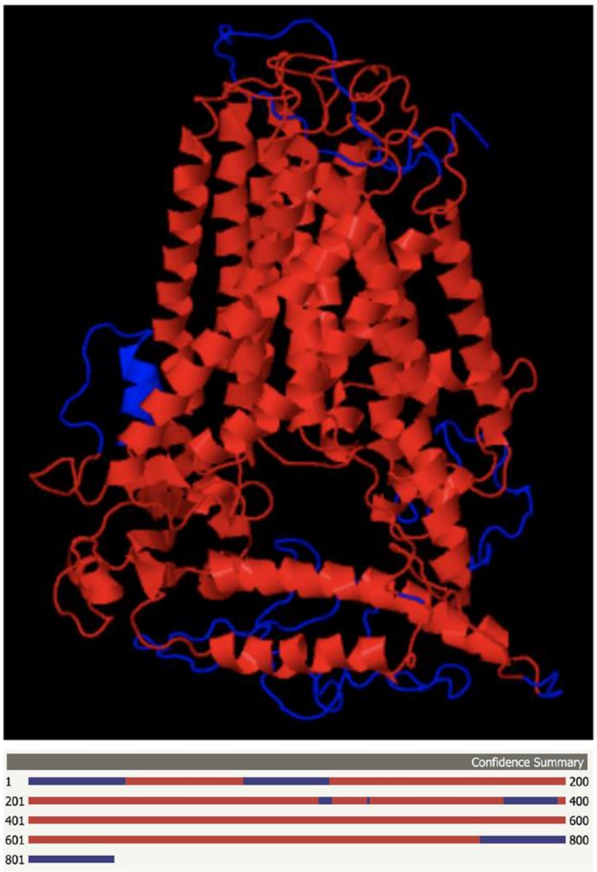

**Supplementary Figure 4 - Projection of confidence score for the TMEM63B protein structure**

In the multi-template homology model obtained by Phyre2, the 81% of residues were modelled with >90% confidence and are showed in red both in the model (upper panel, side vision from the membrane plane) and in the confidence summary (lower panel, linear representation of the amino acids sequence), where low-confidence regions are in blue.

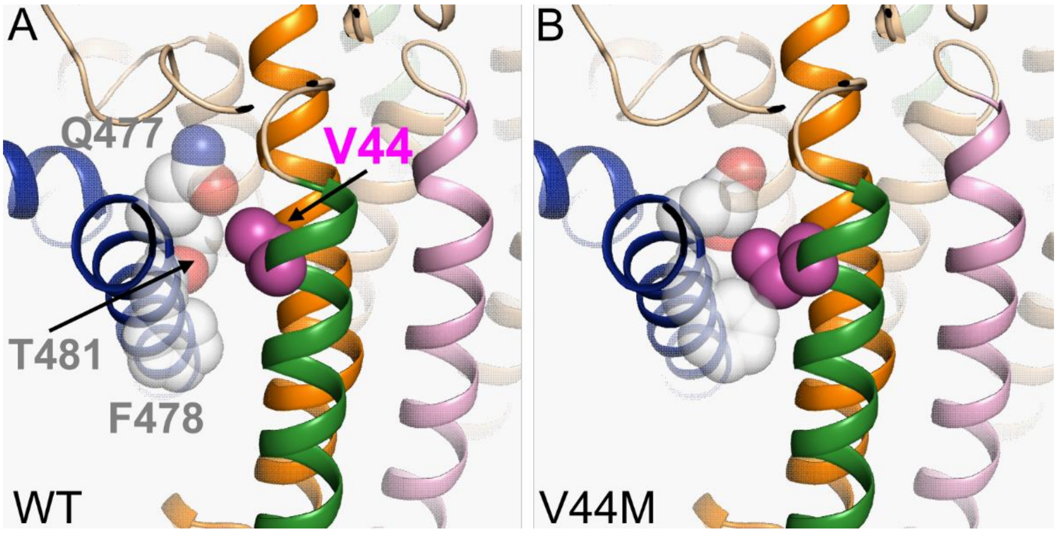

**Supplementary Figure 5 - Close-up of the protein region around Valine 44 in the WT and V44M TMEM63B**

Transmembrane helices are colored as in Figure 3. V44 (purple) and the interacting residues G477, F478, and T481 are indicated by van der Waals spheres in the WT (A) and V44M mutated (B) model of the protein modeled by FoldX.

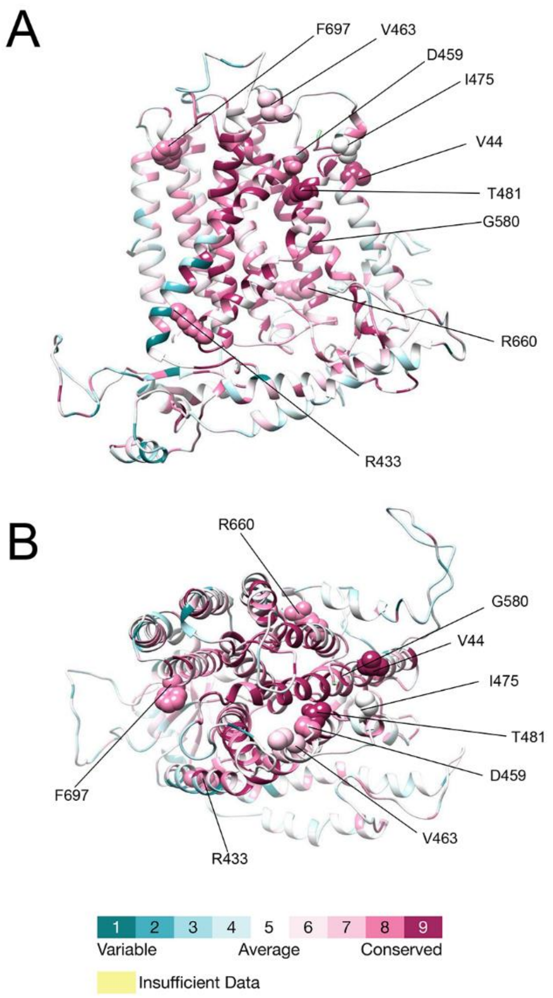

**Supplementary Figure 6 - ConSurf's projection of conservation scores onto the predicted structure of TMEM63B**

View of the predicted structure of TMEM63B from the membrane plane (A) and the extracellular side (B). The color scale is dark aqua (least conserved) to dark magenta (most conserved). Detailed scored and prediction information for each of the residues affected by variants in our cohort are provided in Supplementary Tables 2 and 3.

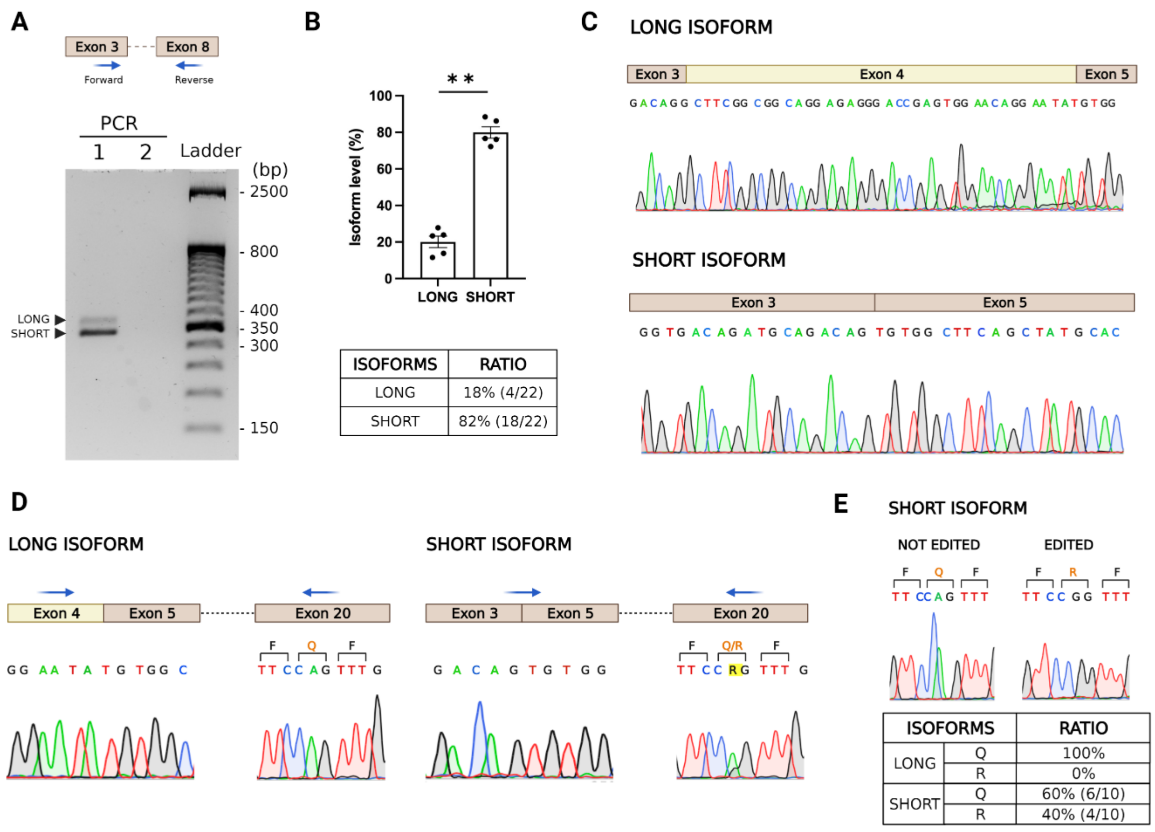

**Supplementary Figure 7 - Characterisation of alternative splicing of exon 4 and Q/R editing at exon 20 in *TMEM63B* RNA from human cerebral cortex**

(A) Upper panel: schematic representation of *TMEM63B* exons 3-8 with arrows indicating Forward and Reverse primers for PCR amplification. Lower panel: agarose gel electrophoresis of the PCR products (Lane 1) showing both long and short *TMEM63B* isoforms, indicated by arrowheads. Lane 2, blank. (B) Upper panel: densitometric quantification of long and short isoforms from gel electrophoresis (data from 5 replicates, expressed as mean ± SEM. **p= 0.0079; Mann-Whitney U test). Lower panel: quantification of long and short isoforms levels obtained from TOPO TA cloning and subsequent sequencing of the PCR products expressed as the number of colonies containing the specific isoform over the total number of screened colonies. (C) Electropherograms from excised bands sequencing showing that exon 4 is only included in the long isoform. (D) Electropherograms of long and short isoforms showing exon 3-5 junctions and editing site at exon 20. A schematic representation of the corresponding exons is indicated, with blue arrows showing Forward and Reverse primers used for PCR amplification. For exon 20 the corresponding amino acids are reported above the nucleotide sequences at the editing site. (E) Upper panel: electropherograms of short edited and not edited isoforms showing the editing site at exon 20, obtained from TOPO TA cloning and subsequent sequencing of the short isoform PCR products. Lower panel: quantification of Q/R editing occurrence in long and short isoforms by TOPO TA technology. For each isoform, the percentage of editing occurrence is obtained from the ratio between the number of colonies edited (R) or not edited (Q) and the total number of analysed colonies.

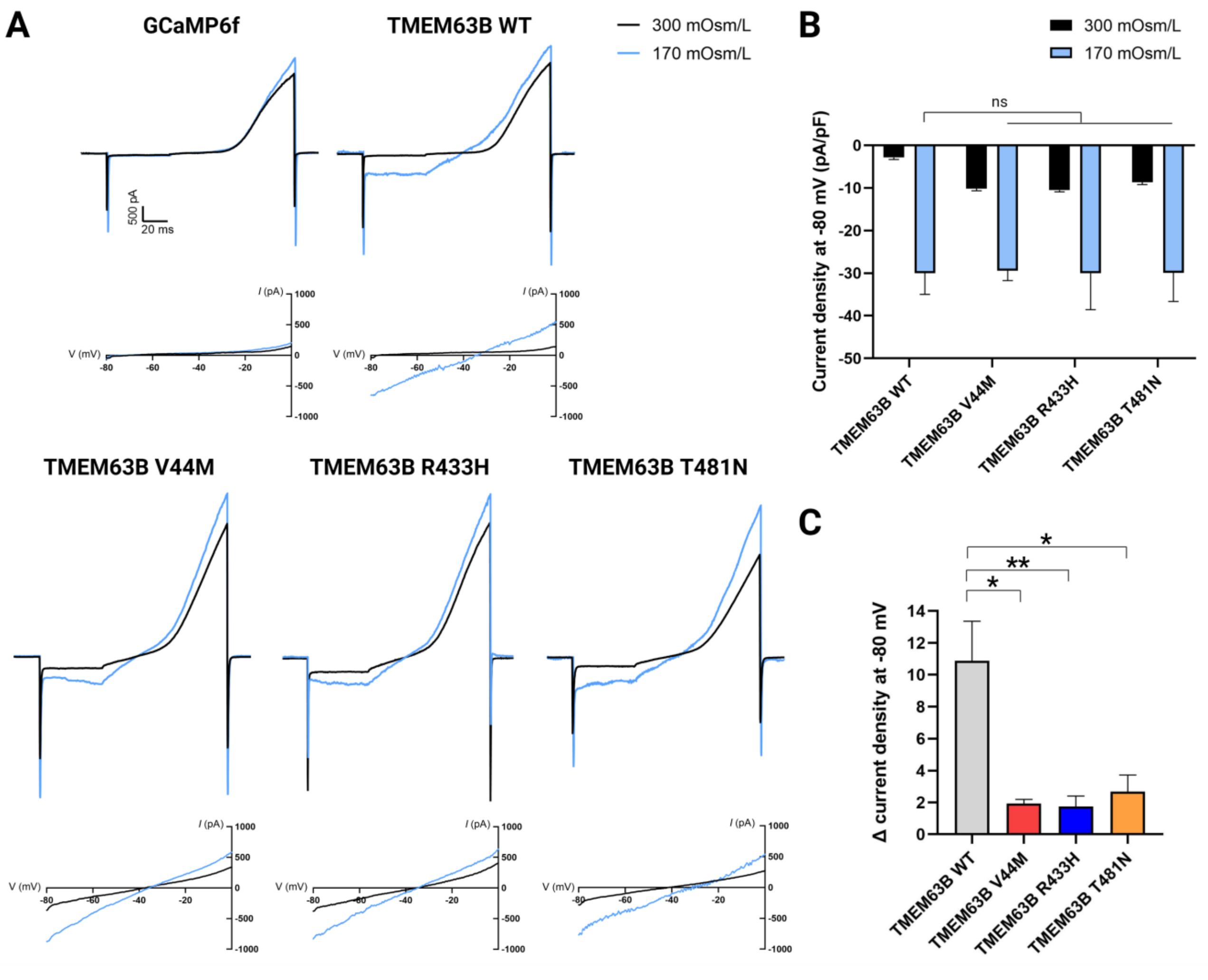

**Supplementary Figure 8 - Effect of hypo-osmotic stimulation on TMEM63B-mediated currents**

(A) Representative current traces and current-voltage (I-V) relationships measured in Neuro2A cells transfected with GCaMP6f, TMEM63B WT or mutant plasmids and recorded with a -80 to + 80mV voltage ramp protocol in isotonic (300 mOsm/L) and hypo-osmotic (170 mOsm/L) conditions. (B) Quantification of whole-cell current density at -80 mV under isotonic and hypo-osmotic conditions (TMEM63B WT, R433H and T481N= 6 cells, TMEM63B V44M= 7 cells, ns= not significant, Kruskal–Wallis and Dunn's multiple comparisons tests). (C) Quantification of the Δ current density measured at -80 mV between hypo-osmotic and isotonic conditions, calculated as:

$$\Delta \text{ current density at -80 mV} = \frac{(\text{hypo-osmotic current density}) - (\text{isotonic current density})}{(\text{isotonic current density})}$$

(TMEM63B WT, R433H and T481N= 6 cells, TMEM63B V44M= 7 cells, **p<0.01, *p<0.05, Kruskal–Wallis and Dunn's multiple comparisons tests).

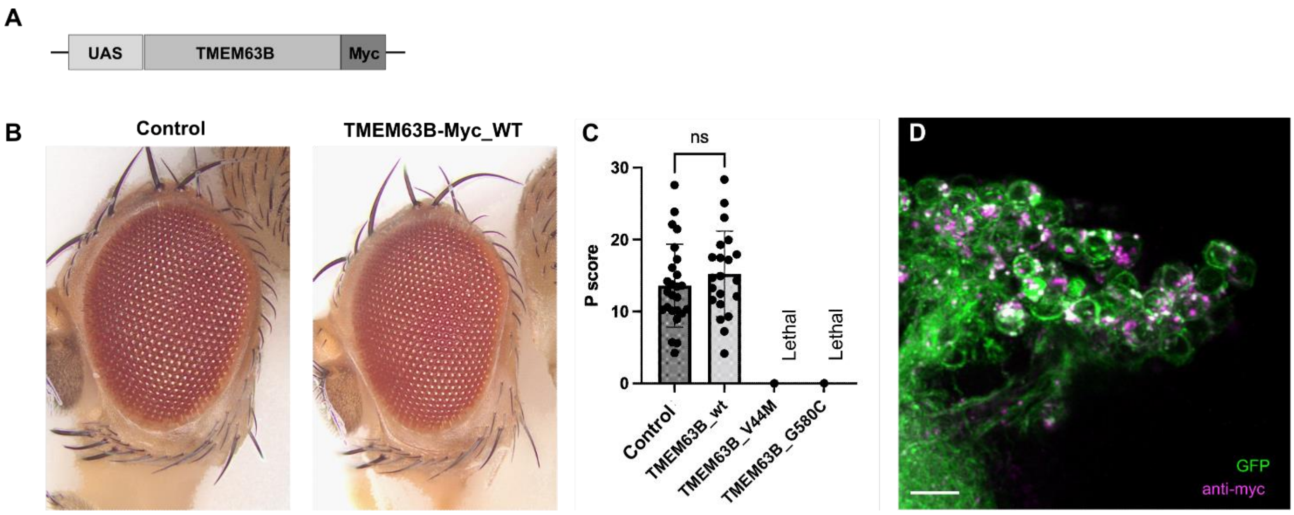

#### Supplementary Figure 9 - Evaluation of human *TMEM63B* variants in *Drosophila*

(A) Cartoon depicting a *TMEM63B* gene construct with a Myc tag in UAS-based vector. (B) Representative bright-field microscope images of the eyes in control and WT TMEM63B-expressing flies (*GMR-Gal4* driver), and (C) quantification of the phenotypic scores in control ($n = 25$) and WT *TMEM63B* ($n = 21$), showing no significant differences in the eye morphology between the control and the WT *TMEM63B*-expressing flies. No data could be obtained for the V44M and G580S-expressing flies, as none of them reached the adult file stage. Data represent the mean ± SD (ns, not significant). (D) Three-dimensional projection of CD8:GFP-labeled Kenyon cells and myc-tagged WT TMEM63B in a posterior brain view in the adult stage. Scale bar = 10 μm.

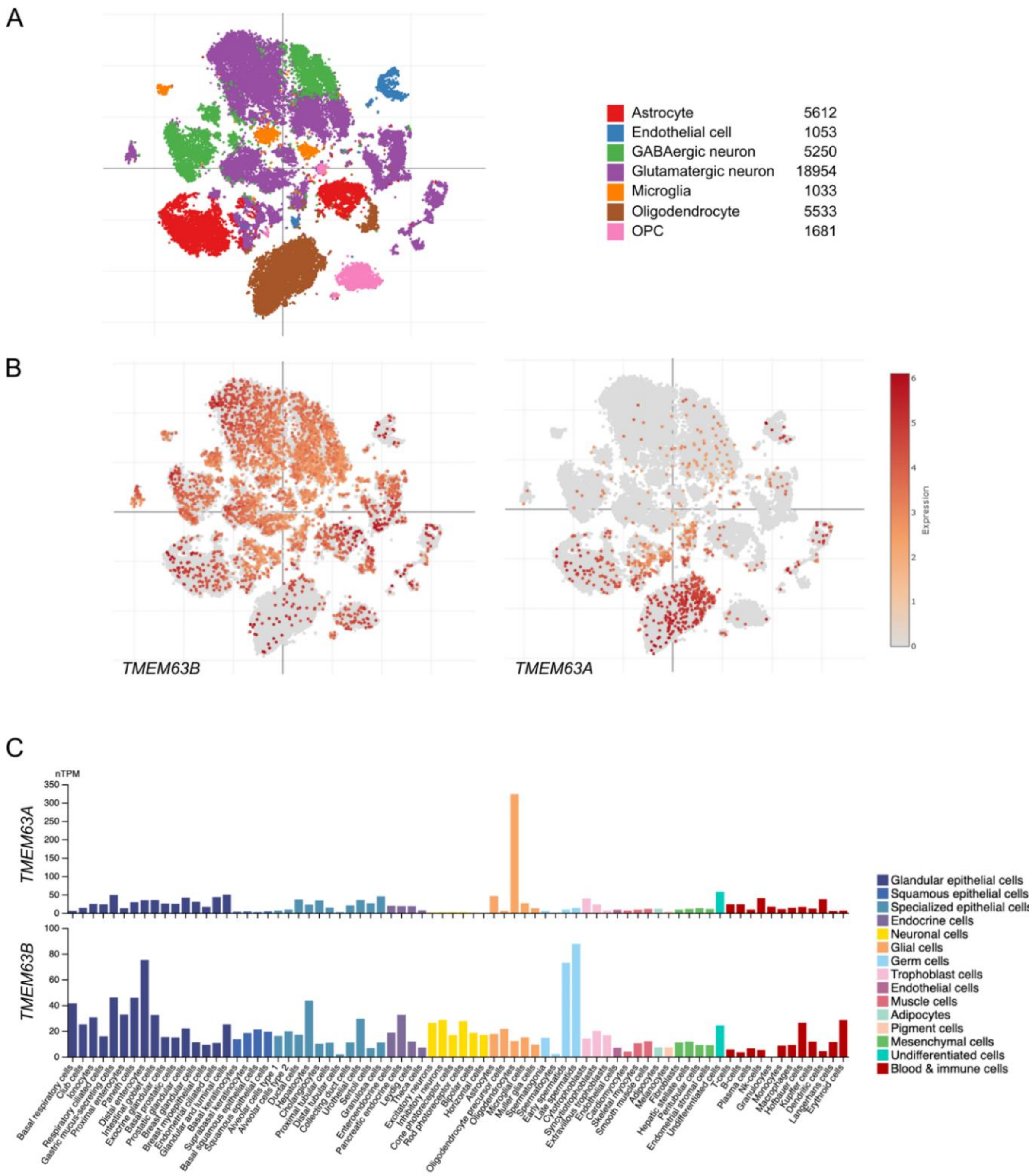

**Supplementary Figure 10 – Single-nucleus expression patterns of *TMEM63B* and *TMEM63A* in the human brain cortex**

Panels A and B show the t-distributed stochastic neighborhood embedding (tSNE) visualization of single-nucleus profiles (dots) colored by cell type (A) and gene expression level (B). Single-nucleus RNA-seq data are shown according to the Single Cell Portal (https://singlecell.broadinstitute.org/single_cell/study/SCP381/) (27) (C) A summary of single cell RNA expression levels of *TMEM63A* and *B* as presented in the Human Protein Atlas (https://www.proteinatlas.org/). Color-coding is based on cell type groups (detailed on the right), each consisting of cell types with functional features in common. Abbreviations: pTPM, transcripts per million.

### Supplementary Tables

#### Supplementary Table 1 – Additional clinical findings of the 16 patients with TMEM63B variants

| Patient number/gender | *TMEM63B* variant (abbreviated form) | Additional clinical findings including haematological manifestations |
|---|---|---|
| **1/M** | V44M | 0-5y: mild anaemia, mildly increased PLT count, stable on yearly FBC; OFT negative |
| **2/M** | R433H | 6-10y: high Hb level; mild abnormalities of RBC, MCV, MCH, no anaemia; OFT negative |
| **3/M** | T481N | None |
| **4/M** | V44M | Jaundice at birth, then intermittent throughout life but less prominent. Macrocytic anaemia (leukoerythroblastic picture with nucleated red cells and myelocytes), borderline monocytosis, hepatosplenomegaly. 0-5y: cholecystectomy for chronic cholecystitis, liver biopsy showed haemosiderin deposits. From 6-10y: persistent macrocytic anaemia with reticulocytosis, increased stomatocytes, iron deficiency anaemia, transfusion dependent (monthly); recurrent chest infections; RCMO onset (flare-up of osteomyelitis associated with Hb reduction and alopecia patches); BM: increased RBC production (due to haemolysis), EMA binding test normal, blood film suggested hereditary elliptocytosis, negative genetic test for hereditary spherocytosis |
| **5/M** | V44M | Jaundice at birth; laryngomalacia (supraglottoplasty at 0-5y); scoliosis |
| **6/F** | V44M | From birth: unexplained macrocytic anaemia, EMA-test analysis normal, haemoglobinopathy screen normal, haemolysis screen negative, negative red cell anaemia TGP, mild hepatosplenomegaly, required transfusion; 0-5y: jaundice; pharyngomalacia and severe OSA leading to respiratory failure and NIV requirement, recurrent chest infections |
| **7/M** | V44M | 0-5y: scleralicterus |
| **8/F** | V44M | 0-5y: severe anaemia, occasional transfusions, episode of haemolysis triggered by infection. 6-10y BM showed myelodysplastic syndrome with aplastic anaemia, haematopoiesis, and chronic haemolysis; scoliosis |
| **9/M** | V44M | Mild and stable anaemia noticed since age 0-5y (no previous measurement available); retinal dystrophy |
| **10/F** | D459E | Fraternal twin, preterm labor, pre-eclampsia, gestational diabetes, pulmonary valve stenosis, hyperbilirubinemia, 0-5y: mild macrocytic anaemia; OFT negative |
| **11/M** | V463I | None |
| **12/F** | I475del | From birth, severe haemolytic anaemia with macrocytosis, thrombocytopenia and hepatosplenomegaly, transfusion dependent, hyperbilirubinemia (total bilirubin 37 µmol/l, direct bilirubin 7 µmol/l), left vesicoureteral reflux (surgical correction 11-15y), chronic urinary retention |
| **13/M** | G580S | None |

| | | |
|---|---|---|
| **14/F** | G580C | Slight bilirubin increase |
| **15/F** | R660T | Birth: mild jaundice, macrocytic anaemia, out-turned feet |
| **16/M** | F697L | None |

Abbreviations and symbols: BM, bone marrow; RCMO, recurrent chronic multifocal osteomyelitis; EMA, eosin-5′-maleimide-binding; F, female; FBC, full blood count; Hb, haemoglobin; HC, head circumference; L, length; M, male; MCH, mean corpuscular haemoglobin; MCV, mean corpuscular volume; NIV, non-invasive ventilation; OFT, osmotic fragility test; OSA, obstructive sleep apnoea; PLT, platelets; RBC, red blood cells; TGP, targeted gene panel; W, weight; y, years.

#### Supplementary Table 2 - Genomic coordinates and in-silico analysis of the *TMEM63B* variants in our cohort

| Pt ID | Variant (genomic GRCh37/hg19) | Protein Change (abbreviation) | Exon | GnomAD (v2.1) | TOPMed (Freeze 8) | CADD | SIFT/score | Polyphen-2/score | MutationTaster/score | PhyloP100way | GERP++ | Grantham score | Tolerance Score (dN/dS, Metadome) |
|---|---|---|---|---|---|---|---|---|---|---|---|---|---|
| 1, 4-9 | 6:g.44102451G>A | V44M | 2 | . | . | 26.6 | deleterious/0 | probably_damaging/0.999 | disease_causing/0.99 | 9.23 | 4.23 | 21 | 0.42 |
| 2 | 6:g.44116567G>A | R433H | 15 | . | . | 29.1 | deleterious/0 | probably_damaging/0.934 | disease_causing/0.99 | 7.73 | 4.23 | 29 | 0.31 |
| 3 | 6:g.44117624C>A | T481N | 16 | . | . | 25.7 | deleterious/0 | probably_damaging/0.992 | disease_causing/1 | 7.59 | 4.48 | 65 | 0.08 |
| 10 | 6:g.44116646C>G | D459E | 15 | . | . | 24.5 | deleterious/0 | probably_damaging/0.943 | disease_causing/0.99 | 3.86 | 4.48 | 45 | 0.15 |
| 11 | 6:g.44116656G>A | V463I | 15 | . | . | 26.5 | deleterious/0 | possibly_damaging/0.85 | disease_causing/0.99 | 9.52 | 4.48 | 29 | 0.16 |
| 12 | 6:g.44117600CCAT>C | I475del | 16 | . | . | NA | NA | NA | disease_causing/1 | 7.31 | 4.48 | NA | 0.11 |
| 13 | 6:g.44119647G>A | G580S | 19 | . | . | 31 | deleterious/0 | probably_damaging/0.999 | disease_causing/1 | 9.58 | 5.01 | 56 | 0.14 |
| 14 | 6:g.44119647G>T | G580C | 19 | . | . | 32 | deleterious/0 | probably_damaging/1 | disease_causing/1 | 10 | 5.01 | 159 | 0.14 |
| 15 | 6:g.44121449G>C | R660T | 21 | . | . | 26.6 | deleterious/0 | probably_damaging/0.955 | disease_causing/0.99 | 9.26 | 4.59 | 71 | 0.31 |
| 16 | 6:g.44121559T>C | F697L | 21 | . | . | 30 | deleterious/0 | probably_damaging/0.968 | disease_causing/0.99 | 7.52 | 4.62 | 22 | 0.06 |

The table shows the coordinates of the variants according to the recommendations of the Human Genome Variation Society (http://varnomen.hgvs.org/), based on the hGRCh37/hg19 assembly and the NM_018426.3 reference transcript. None of the variants was reported in publicly available allele frequency databases such as GnomAD (v2.1) and TOPMed (Freeze 8). For all the variants in our cohort we report the *in-silico* predictions of pathogenicity obtained from multiple tools: for CADD (Combined Annotation-

Dependent Depletion) the PHRED-like scaled C-score greater or equal 20 indicates the 1% most deleterious substitutions to the gene products (28); SIFT confidence score for a missense variant is computed as 1-p where p is the probability for the variant to be deleterious (29); for MutationTaster a value close to 1 indicates a high 'security' of the prediction, while for Polyphen-2 it represents the probability for the variant to be disease causing in a 0-1 range) (30). PhyloP100way and GERP++ scores range from -14.1 to 6.4 and -12.3 to 6.17 respectively, with higher scores indicating stronger constraint (5, 6). Grantham score ranges from 5 to 215 and predicts the distance between two amino acids, in an evolutionary sense. Higher Grantham scores are considered more deleterious (31). Tolerance Scores (dN/dS nonsynonymous over synonymous ratio according to Metadome) range from highly intolerant (0-0.19) to intolerant (0.2-0.49) (7). NA: not available/not applicable.

**Supplementary Table 3 – Structural analysis of the *TMEM63B* variants by Consurf and Missense3D**

| Pt ID | Protein Change | Transmembrane domain | Protein domain (Pfam 35.0) | ConSurf scale/prediction | Missense3D prediction |
|---|---|---|---|---|---|
| 1, 4-9 | V44M | TM1 | NA | 9/predicted structural residue (highly conserved and buried) | Expansion of cavity volume by 8.208 Å^3 |
| 2 | R433H | TM4 | PF02714 | 8/buried residue | Contraction of cavity volume by 3.888 Å^3 |
| 3 | T481N | TM5 | PF02714 | 9/predicted structural residue (highly conserved and buried) | No structural damage detected; Unable to calculate cavity in mutant structure |
| 10 | D459E | TM4 | PF02714 | 8/predicted functional residue (highly conserved and exposed) | Buried salt bridge breakage (Asp 459, LYS 460) |
| 11 | V463I | TM4 | PF02714 | 8/buried residue | Contraction of cavity volume by 13.824 Å^3 |
| 12 | I475del | TM5 | PF02714 | 5/buried residue | NA |
| 13 | G580S | TM7 | PF02714 | 9/predicted structural residue (highly conserved and buried) | Buried Gly residue (RSA 5.9%) replaced with a buried Ser residue (RSA 3.8%); Contraction of cavity volume by 52.056 Å^3 |
| 14 | G580C | TM7 | PF02714 | 9/predicted structural residue (highly conserved and buried) | Buried Gly residue (RSA 5.9%) replaced with a buried Cys residue (RSA 3.7%); Contraction of cavity volume by 66.744 Å^3 |
| 15 | R660T | TM8 | PF02714 | 8/predicted functional residue (highly conserved and exposed) | Buried charge replaced (Arg, RSA 5.6%) with an uncharged residue (Thr); Buried salt bridge breakage (Arg 660, Asp 137); Expansion of cavity volume by 60.696 Å^3 |
| 16 | F697L | TM9 | PF02714 | 8/buried residue | Expansion of cavity volume by 30.24 Å^3 |

All the affected residues map in a transmembrane (TM) domain, and all but the recurrent V44M are in the RSN1_7TM (PF02714) domain (32). For all the affected residues, we report the conservation scale (ranging from variable, 1 to conserved, 9) and the neural-network algorithm prediction from ConSurf (https://consurf.tau.ac.il/) (15). For all missense variants, we also show the prediction of possible structural changes according to Missense3D-DB, which considered 17 structural features, including secondary structure alterations, non-covalent bond breakages, and buried residues changes (16). NA: not available/not applicable.

**Supplementary Table 4 – Methods for exome/genome sequencing in the cohort.**

| Pt ID | Sequencing approach, with reference |
|---|---|
| **1-3** | trio WES (33) |
| **4** | trio WGS (34, 35) |
| **5** | trio WES (36) |
| **6** | trio WGS (37) |
| **7** | singleton WES (38) |
| **8, 14** | trio WES (39) |
| **9** | trio WES (40) |
| **10** | singleton WES (41) |
| **11** | trio WES (42) |
| **12** | trio WES (43) |
| **13** | trio WES (44) |
| **15** | trio WES (45) |
| **16** | trio WES (46) |

#### Study Groups

****TMEM63B collaborators:** Francesca Pochiero[1], Francesco Mari[1], Venkateswaran Ramesh [2], Valeria Capra[3], Margherita Mancardi[3], Boris Keren[4], Cyiril Mignot[4,5], Matteo Lulli[6], Kendall Parks[7], Helen Griffin[8], Melanie Brugger[9], Telethon Undiagnosed Diseases Program (TUDP) consortium[10], Vincenzo Nigro[10,11], Yuko Hirata[12], Reiko Koichihara[12], Borut Peterlin[13], Yuko Hirata[14], Ryuto Maki[15], Yohei Nitta[16]

1. Neuroscience Department, Meyer Children's Hospital, Florence, Italy
2. Department of Paediatric Neurology, Newcastle upon Tyne Hospitals NHS Trust, Newcastle upon Tyne, UK
3. Department of Neurosciences, Rehabilitation, Ophthalmology, Genetics, Maternal and Child Health, University of Genoa, Genoa, Italy
4. Département de Génétique and Centre de Référence Déficiences Intellectuelles de Causes Rares, AP-HP.Sorbonne Université, Hôpital Pitié-Salpêtrière, Paris, France
5. INSERM, U 1127, CNRS UMR 7225, Sorbonne Universités, UPMC Univ Paris 06 UMR S 1127, Institut du Cerveau, ICM, Paris, France
6. Department of Experimental and Clinical Biomedical Sciences "Mario Serio", University of Florence, Florence, Italy
7. Department of Neurology and Institute of Human Genetics and Weill Institute for Neurosciences, University of California, San Francisco, San Francisco, CA, USA
8. Newcastle University Translational and Clinical Research Institute, Newcastle upon Tyne, UK
9. Institute of Human Genetics, School of Medicine, Technical University Munich, Munich, Germany
10. Telethon Institute of Genetics and Medicine (TIGEM), Pozzuoli, Italy
11. Department of Precision Medicine, University "Luigi Vanvitelli", Naples, Italy
12. Division of Neurology, Saitama Children's Medical Center, Saitama, Japan
13. Clinical Institute of Genomic Medicine, University Medical Center Ljubljana, Ljubljana, Slovenia
14. Division of Neurology, Saitama Children's Medical Center, Saitama, Japan
15. School of Life Science and Technology, Tokyo Institute of Technology, Yokohama, Kanagawa, Japan
16. Brain Research Institute, Niigata University, Niigata 951-8585, Japan

**§The Genomics England Research Consortium:** J. C. Ambrose[1], P. Arumugam[1], R. Bevers[1], M. Bleda[1], F. Boardman-Pretty[1,2], C. R. Boustred[1], H. Brittain[1], M. A. Brown[1], M. J. Caulfield[1,2], G. C. Chan[1], A. Giess[1], J. Griffin[1], A. Hamblin[1], S. Henderson[1,2], T. J. P. Hubbard[1], R. Jackson[1], L. J. Jones[1,2], D. Kasperaviciute[1,2], M. Kayikci[1], A. Kousathanas[1], L. Lahnstein[1], A. Lakey[1], S. E. A Leigh[1], I. U. S. Leong[1], F. J. Lopez[1], F. Maleady-Crowe[1], M. McEntagart[1], F. Minneci[1], J. Mitchell[1], L. Moutsianas[1,2], M. Mueller[1,2], N. Murugaesu[1], A. C. Need[1,2], P. O'Donovan[1], C. A. Odhams[1], C. Patch[1,2], D. Perez-Gil[1], M. B. Pereira[1], J. Pullinger[1], T. Rahim[1], A. Rendon[1], T. Rogers[1], K. Savage[1], K. Sawant[1], R. H. Scott[1], A. Siddiq[1], A. Sieghart[1], S. C. Smith[1], A. Sosinsky[1,2], A. Stuckey[1], M. Tanguy[1], A. L. Taylor Tavares[1], E. R. A. Thomas[1,2], S. R. Thompson[1], A. Tucci[1,2], M. J. Welland[1], E. Williams[1], K. Witkowska[1,2], S. M. Wood[1,2], M. Zarowiecki[1]

1. Genomics England, London, UK
2. William Harvey Research Institute, Queen Mary University of London, London, EC1M 6BQ, UK

28. Rentzsch P, Witten D, Cooper GM, Shendure J, Kircher M. CADD: predicting the deleteriousness of

variants throughout the human genome.. *Nucleic Acids Res.* 2019;47(D1):D886–D894.
